# Whole-genome sequencing analysis of left ventricular structure and sphericity in 80,000 people

**DOI:** 10.1101/2025.08.22.25334019

**Authors:** James P. Pirruccello

## Abstract

**Background:** Sphericity is a measurement of how closely an object approximates a globe. The sphericity of the blood pool of the left ventricle (LV), is an emerging measure linked to myocardial dysfunction.

**Methods:** Video-based deep learning models were trained for semantic segmentation (pixel labeling) in cardiac magnetic resonance imaging in 84,327 UK Biobank participants. These labeled pixels were co-oriented in 3D and used to construct surface meshes. LV ejection fraction, mass, volume, surface area, and sphericity were calculated. Epidemiologic and genetic analyses were conducted. Polygenic score validation was performed in *All of Us*.

**Results:** 3D LV sphericity was found to be more strongly associated (HR 10.3 per SD, 95% CI 6.1-17.3) than LV ejection fraction (HR 2.9 per SD reduction, 95% CI 2.4-3.6) with dilated cardiomyopathy (DCM). Paired with whole genome sequencing, these measurements linked LV structure and function to 366 distinct common and low-frequency genetic loci—and 17 genes with rare variant burden—spanning a 25-fold range of effect size. The discoveries included 22 out of the 26 loci that were recently associated with DCM. LV genome-wide polygenic scores were equivalent to, or outperformed, dedicated hypertrophic cardiomyopathy (HCM) and DCM polygenic scores for disease prediction. In *All of Us*, those in the polygenic extreme 1% had an estimated 6.6% risk of DCM by age 80, compared to 33% for carriers of rare truncating variants in the gene *TTN*.

**Conclusions:** 3D sphericity is a distinct, heritable LV measurement that is intricately linked to risk for HCM and DCM. The genetic findings from this study raise the possibility that the majority of common genetic loci that will be discovered in future large-scale DCM analyses are present in the current results.

## Introduction

The failing heart takes on a spherical shape. For decades, an increased left ventricular (LV) sphericity (sometimes referred to as *globularity*) has been reported in patients with myocardial dysfunction^1^ and regurgitant lesions of the aortic^2^ and mitral valves^3,4^. When formalized as a quantitative trait, greater LV sphericity has also been linked to atrial fibrillation, heart failure, and cardiomyopathy^5,6^.

The heritability of standard measurements of LV structure and function have become well established: dozens of genetic loci have now been linked to LV measurements through genome-wide association studies (GWAS)^7,8^. There is also emerging evidence that LV sphericity itself is a heritable trait: Vukadinovic, *et al*, previously linked the LV aspect ratio at end-diastole to four genetic loci (near *PDZRN3*, *HLA*, *PLN*, and *ANGPT1*)^6^. Of these, *PDZRN3* was also among the 26 dilated cardiomyopathy (DCM)-linked loci in the recent HERMES2.0 DCM GWAS^9^. These findings suggest a genetic relationship between LV sphericity and DCM, but data on the genetic underpinnings of sphericity remain sparse.

The present work introduces a 3D surface mesh-based measurement for sphericity to capture the LV’s geometry more comprehensively than low-dimensional representations, such as the aspect ratio^6^ or the length-volume method^4,5^. In contrast to prior work that has examined sphericity in diastole, this work also measures sphericity in systole, when the LV is maximally contracted. These data enable the largest genetic analysis of LV structure and function to date in over 80,000 UK Biobank participants.

## Results

Analyses were conducted in 84,327 participants from the UK Biobank magnetic resonance imaging (MRI) substudy (**Methods**; **Supplementary Figure 1**)^10,11^. Rare variant and polygenic score findings were validated in *All of Us*^12^.

### Measuring left ventricular structure and sphericity

Deep learning video segmentation models were trained to identify the LV blood pool and myocardial pixels in UK Biobank cardiac MRI (**Methods**), achieving Dice scores in the test set from 0.94-0.96 for the blood pool and 0.83-0.88 for the myocardium (1.0 being a perfect score)^13,14^. The short-axis (SAX) segmentation model was externally validated in the ACDC dataset^15^, where it achieved a Dice score of 0.92 for the blood pool and 0.83 for the myocardium (**Supplementary Table 1**).

The trained models were applied to MRI data from 84,327 UK Biobank participants (**Figure 1**). Segmented cardiac pixels were co-registered in 3D space based on DICOM metadata and used to construct surface meshes^16,17^. After technical quality control (**Supplementary Methods**), LV measurements from 82,674 participants were retained for downstream analyses (**Table 1**, **Supplementary Figure 2**).

**Figure 1:**
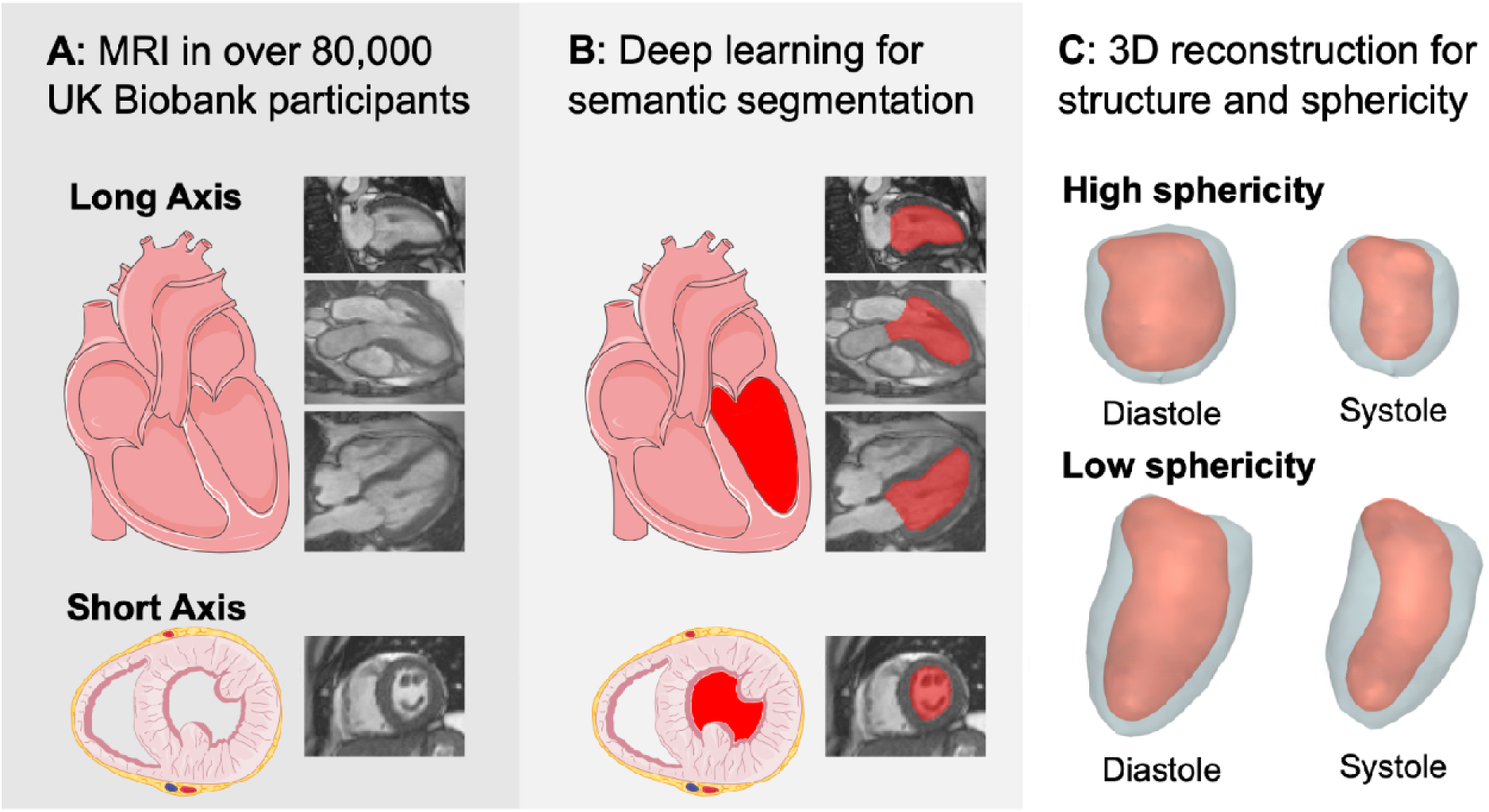
MRI, semantic segmentation, and 3D reconstruction. **Panel A**: Long-axis (top) and short-axis (bottom) MRI still frames. **Panel B**: The MRI still frames have been overlaid with a red-colored tint based on the output from the deep learning semantic segmentation model that identified the pixels belonging to the left ventricular blood pool. **Panel C**: Rendering of 3D LV reconstructions with high sphericity (top) and low sphericity (bottom). The blood pool is tinted red, and the myocardium light gray. The images are oriented with the left ventricular outflow tract at the top left. The heart drawings were provided by *Servier Medical Art*, licensed under *CC BY 4.0*. The magnetic resonance images were reproduced by kind permission of UK Biobank ©.

**Table 1:**
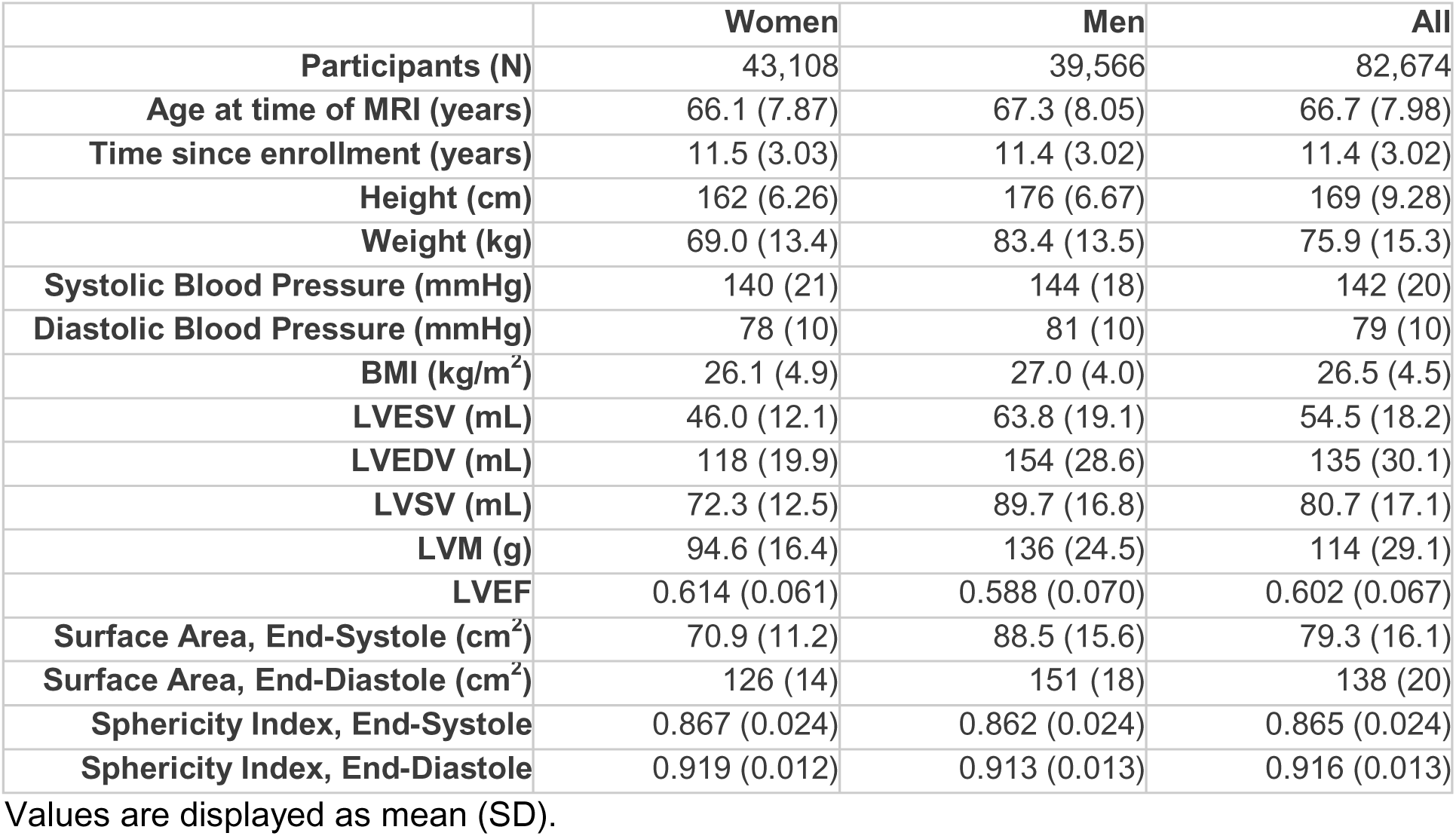
Population characteristics.

|  | Women | Men | All |
| --- | --- | --- | --- |
| **Participants (N)** | 43,108 | 39,566 | 82,674 |
| **Age at time of MRI (years)** | 66.1 (7.87) | 67.3 (8.05) | 66.7 (7.98) |
| **Time since enrollment (years)** | 11.5 (3.03) | 11.4 (3.02) | 11.4 (3.02) |
| **Height (cm)** | 162 (6.26) | 176 (6.67) | 169 (9.28) |
| **Weight (kg)** | 69.0 (13.4) | 83.4 (13.5) | 75.9 (15.3) |
| **Systolic Blood Pressure (mmHg)** | 140 (21) | 144 (18) | 142 (20) |
| **Diastolic Blood Pressure (mmHg)** | 78 (10) | 81 (10) | 79 (10) |
| **BMI (kg/m<sup>2</sup>)** | 26.1 (4.9) | 27.0 (4.0) | 26.5 (4.5) |
| **LVESV (mL)** | 46.0 (12.1) | 63.8 (19.1) | 54.5 (18.2) |
| **LVEDV (mL)** | 118 (19.9) | 154 (28.6) | 135 (30.1) |
| **LVSV (mL)** | 72.3 (12.5) | 89.7 (16.8) | 80.7 (17.1) |
| **LVM (g)** | 94.6 (16.4) | 136 (24.5) | 114 (29.1) |
| **LVEF** | 0.614 (0.061) | 0.588 (0.070) | 0.602 (0.067) |
| **Surface Area, End-Systole (cm<sup>2</sup>)** | 70.9 (11.2) | 88.5 (15.6) | 79.3 (16.1) |
| **Surface Area, End-Diastole (cm<sup>2</sup>)** | 126 (14) | 151 (18) | 138 (20) |
| **Sphericity Index, End-Systole** | 0.867 (0.024) | 0.862 (0.024) | 0.865 (0.024) |
| **Sphericity Index, End-Diastole** | 0.919 (0.012) | 0.913 (0.013) | 0.916 (0.013) |

Values are displayed as mean (SD).

The sphericity index is a value ranging from 0-1, with 1 representing a sphere (**Supplementary Table 2**)^18^. The mean LV sphericity index at end-diastole (the moment of maximum LV filling; “sphericity-ED”) was 0.916 (SD 0.013), while that for sphericity at end-systole (maximum LV contraction; “sphericity-ES”) was 0.865 (SD 0.024; **Table 1**; **Supplementary Figure 3**). While all LV measurements deviated to a measurable degree from a Gaussian distribution, the sphericity measurements deviated by a smaller degree than any of the standard LV measurements other than LV ejection fraction (LVEF), as measured by the earth mover’s distance (0.100 standard deviation [SD] for sphericity-ES, 0.065 SD for sphericity-ED, and 0.034 SD for LVEF; **Supplementary Methods**; **Supplementary Table 3**).

Sphericity was only partially predictable from the standard LV measurements: LVEF, LV end diastolic volume (LVEDV), LV end systolic volume (LVESV), LV mass (LVM), and LV stroke volume (LVSV), along with age and sex, accounted for 22% of the variation in sphericity-ED and 56% in sphericity-ES (**Supplementary Figure 4**). Sphericity-ES was more independent of sex (r=0.10) and height (r=0.00) than were the standard LV measurements (**Supplementary Figure 5**).

### Opposing associations with dilated and hypertrophic cardiomyopathies

Individuals with a diagnosis of DCM prior to imaging had greater sphericity-ES compared to unaffected individuals (N=53, +1.58 SD, P=1.5E-31). Those with hypertrophic cardiomyopathy (HCM) had lower sphericity-ES (N=47, -0.81 SD, P=1.4E-08). This opposing pattern was unique to sphericity: LVEF and LVESV were predominantly abnormal in DCM, while LVM was elevated in both cardiomyopathies (**Figure 2**; **Supplementary Table 4**).

**Figure 2:**
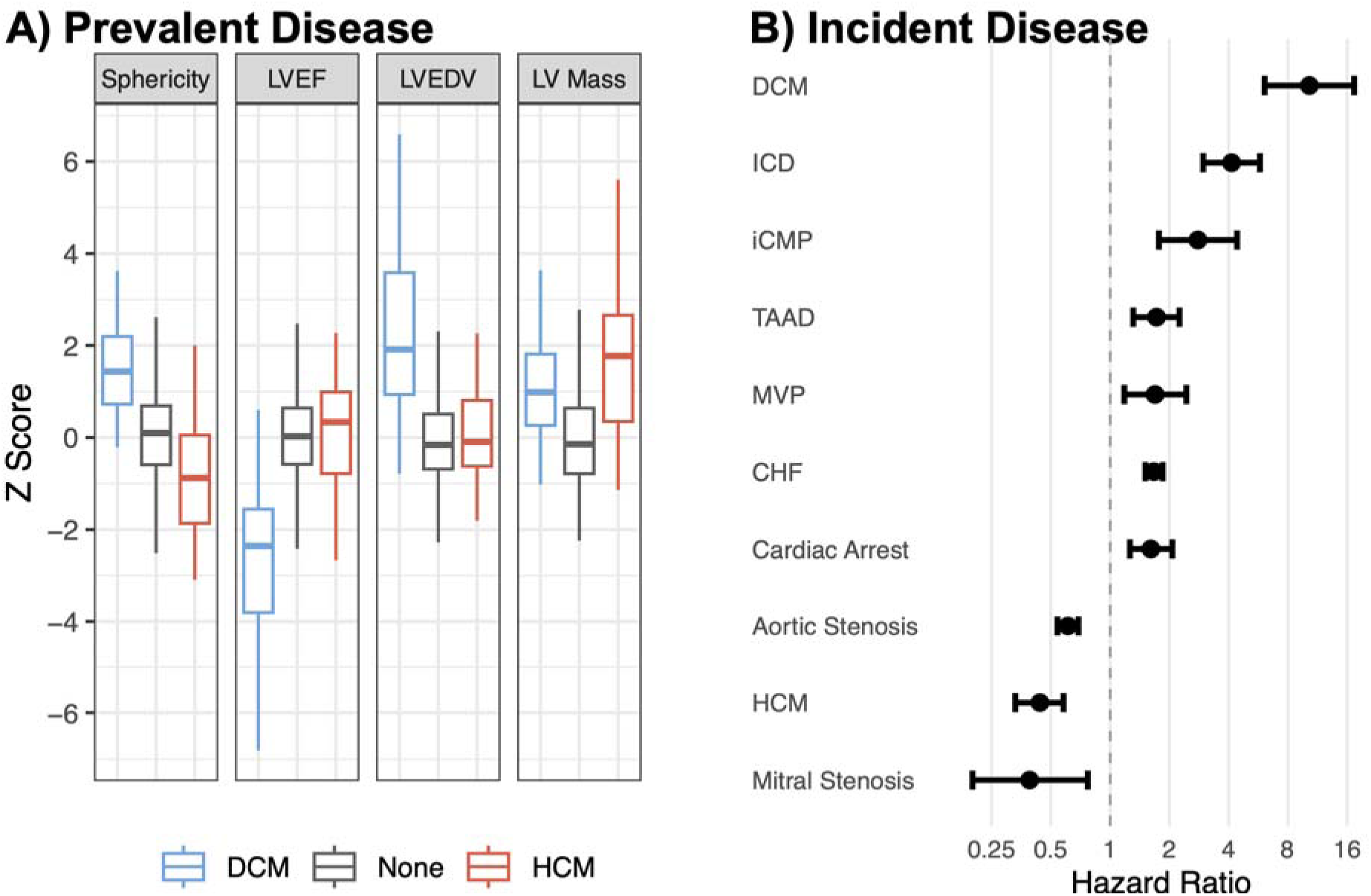
LV sphericity and cardiomyopathies. **Panel A**: Association between LV measurements and DCM or HCM present prior to imaging. Sphericity: sphericity-ES; LVEF: LV ejection fraction; LVEDV: LV end diastolic volume. **Panel B**: Hazard ratios for incident disease occurring after the imaging visit. ICD: implantable cardioverter-defibrillator implantation; iCMP: ischemic cardiomyopathy; TAAD: thoracic aortic aneurysm and dissection; MVP: mitral valve prolapse; CHF: congestive heart failure.

Among individuals who had no previous diagnosis of DCM, greater LV sphericity-ES conferred a hazard ratio (HR) per SD of 10.3 for incident DCM (95% confidence interval [CI] 6.1-17.3) during a mean of 3.9 years of follow-up after MRI (**Figure 2**). LV sphericity-ED had a more modest HR per SD of 3.5 (95% CI 2.1-5.6). When applied in the same participants, both point estimates exceeded the effect size from computing length-volume-based sphericity (HR 2.0 per SD, 95% CI 1.6-2.7)^4,5^. Further, these relationships were greater in magnitude than those of the standard LV measurements such as LVEF (HR 2.9 per SD reduction, 95% CI 2.4-3.6). When tested together in a joint model, both LVEF and sphericity-ES remained significantly associated with DCM (**Supplementary Figure 6**). This pattern was also seen for diagnoses based on procedural codes, such as implantable cardioverter-defibrillator implantation (HR 4.1 per SD of sphericity-ES [95% CI 3.0-5.8], compared to HR 2.6 per SD decrease of LVEF [95% CI 2.3-3.0]).

Conversely, lower sphericity-ES conferred a 2.3-fold higher hazard for HCM (HR 2.3 per SD reduction, 95% CI 1.7-3.0), whereas the length-volume-based sphericity measurement was unassociated (HR 1.1 per SD, 95% CI 0.7-1.5). While weaker than the relationship between HCM and LV mass (HR 2.7, 95% CI 2.3-3.2), both LV mass and sphericity-ES remained significantly associated with HCM in a joint model (**Supplementary Figure 6**).

Sphericity was also associated with subsequent diagnoses of aortic stenosis, mitral valve prolapse, mitral stenosis, heart failure, and cardiac arrest (**Figure 2**; **Supplementary Table 5**). Sphericity was not significantly associated with all-cause mortality overall; however, there was a nominal interaction with heart failure (717 heart failure cases, 807 deaths, 27 deaths among heart failure cases, HR 1.36 per SD of sphericity-ES among those with heart failure, interaction P=0.035).

### GWAS of left ventricular structure and sphericity

Data from 80,389 participants passed quality control (80,113 with valid sphericity measurements) and contributed to genetic analyses (**Supplementary Figure 1**). LV sphericity had BOLT-REML heritability estimates (0.36 for sphericity at ED and 0.35 at ES) that were intermediate between those for LVEF (0.31) and LVEDV (0.40; **Supplementary Table 6**).

Compared to LVEF, which is also dimensionless, LV sphericity was less genetically correlated with chamber size (*rg* with LVESV was -0.82 for LVEF and 0.69 for LV sphericity-ES; **Supplementary Table 7**, **Supplementary Figure 7**). The GWAS *ldsc* intercepts ranged from 0.98 for sphericity-ED to 1.04 for LVM (**Supplementary Table 8**), consistent with polygenicity^19^.

In the primary analysis, GWAS was conducted for nine LV traits, yielding 319 distinct loci with P<5E-08 (834 locus-trait pairs): 95 for LV sphericity-ED, 100 for LV sphericity-ES (**Figure 3**), 94 for LVEDV, 118 for LVESV, 50 for LVSV, 95 for LVEF, 72 for LV mass, 92 for surface area at ED, and 118 for surface area at ES (**Supplementary Figure 8**; **Supplementary Table 9**). 222 distinct loci were associated with the five standard LV measurements (LVEDV, LVESV, LVEF, SV, and LVM). 87 additional loci were distinct to the sphericity GWAS (**Table 2**; **Supplementary Figure 9**) and 10 additional distinct loci were associated with LV surface area. Sensitivity analyses that indexed the phenotypes to body surface area yielded an additional 47 distinct loci (**Supplementary Table 9**) for a total of 366 distinct loci (1,115 locus-trait pairs) across the 13 GWAS. 10 distinct loci harbored low-frequency single-nucleotide variants (MAF 0.001-0.01; **Table 3**), including missense variants in *RBM20*, *FLNC*, *CSRP3*, and *ZFAT* (**Figure 4**). Two loci (near *TTN* and *TBX3*) were associated with P<5E-08 for all 13 LV measurements. Nine loci were identified on chromosome X (near *DMD*, *DDX3D*, *DIPK2B*, *ZNF674*, *GPKOW*, *DGKK*, *ITIH6*, *ATRX*, and *SEPTIN6*). Of note, *DMD* encodes dystrophin, which forms the dystrophin glycoprotein complex in combination with dystroglycan, sarcospan, syntrophin, and laminin (all encoded by the nearest gene to a lead variant from the present GWAS) as well as sarcoglycan, dystrobrevin, caveolin-3, and neuronal nitric oxide synthase^20^.

**Figure 3:**
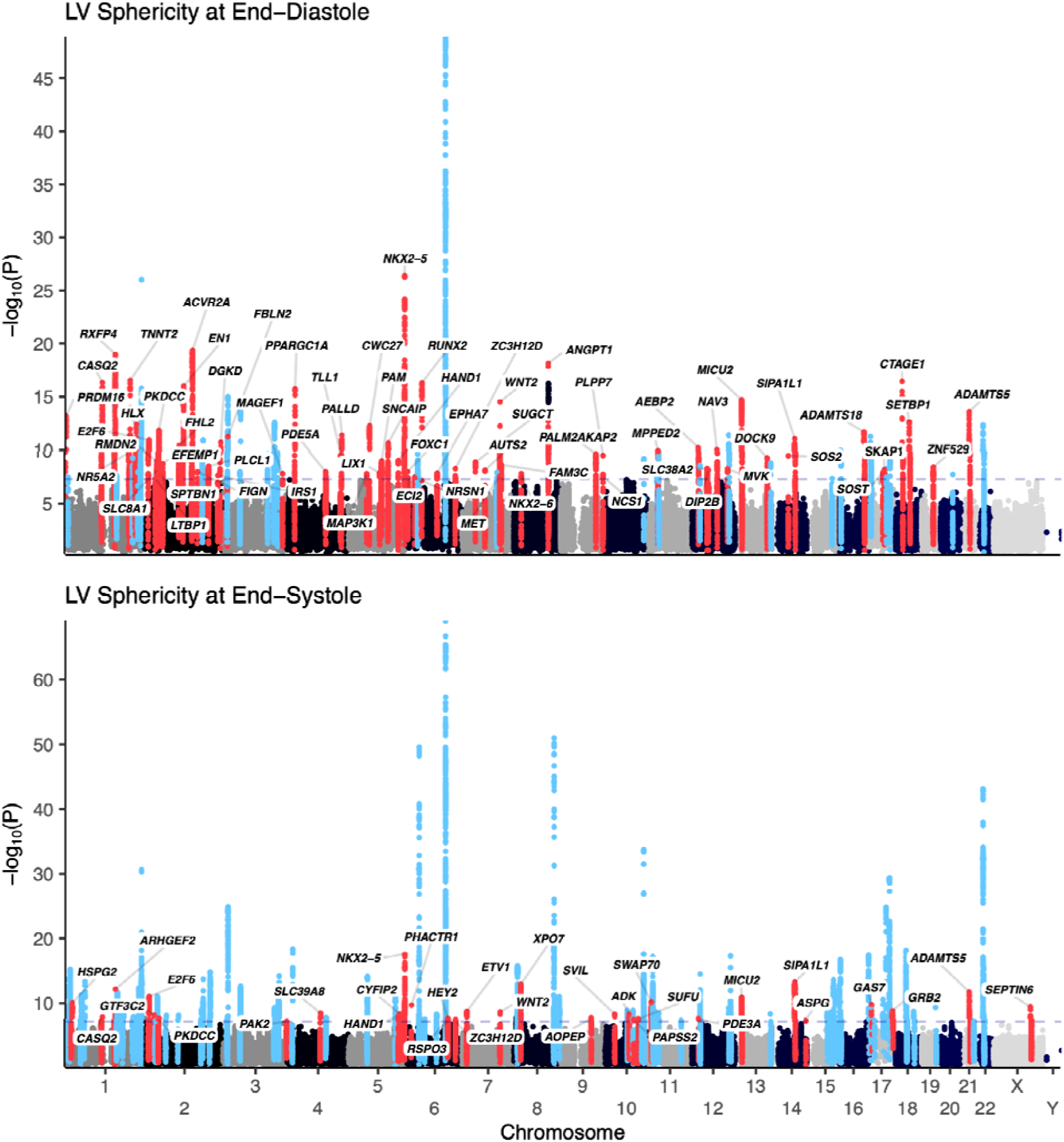
Manhattan plot for sphericity. Manhattan plots for sphericity-ED and sphericity-ES are depicted. Blue-tinted loci have a lead variant with P<5E-08 for sphericity and for the standard LV measurements. Red-tinted loci have a lead variant with P<5E-08 for sphericity and not for the standard LV measurements; for these loci, the gene nearest to the lead variant is labeled.

**Figure 4:**
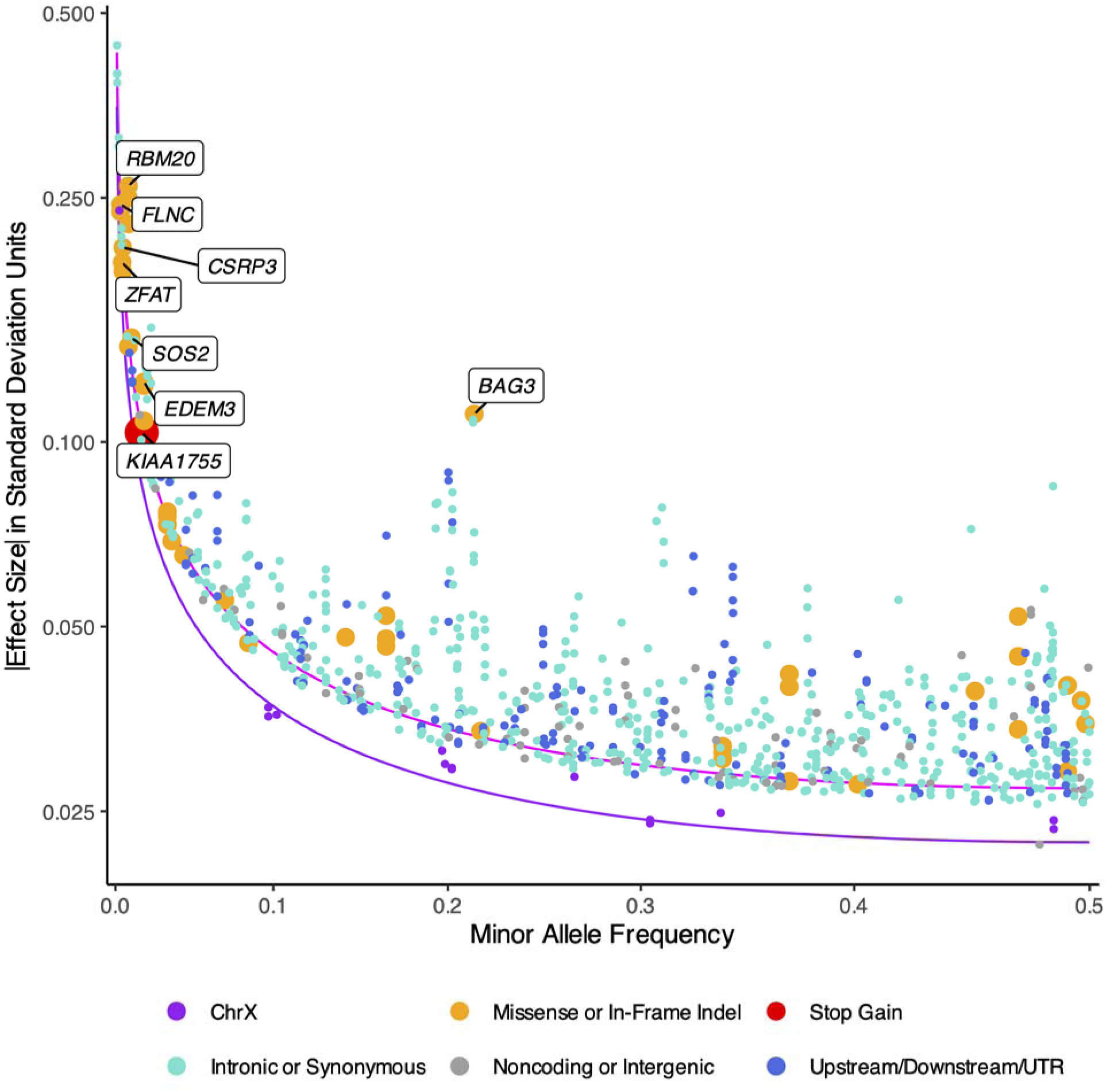
Effect sizes across the allele frequency spectrum. Independent variants for the primary and secondary GWAS are displayed by minor allele frequency (**x-axis**) and absolute value of the effect estimate (**y-axis**). All available independent variants with MAF ≥ 0.001 were tested. Variants are colorized based on functional groups or presence on chromosome X. Missense and nonsense variants are displayed in a larger point size, and those with effect estimates greater than 0.1 SD additionally have a label (the name of the nearest gene) applied to their first occurrence. An approximate 50% power frontier at α=5•10^-8^ for N=80,000 is displayed in magenta for autosomes and in purple for chromosome X (**Supplementary Methods**).

**Table 2:**
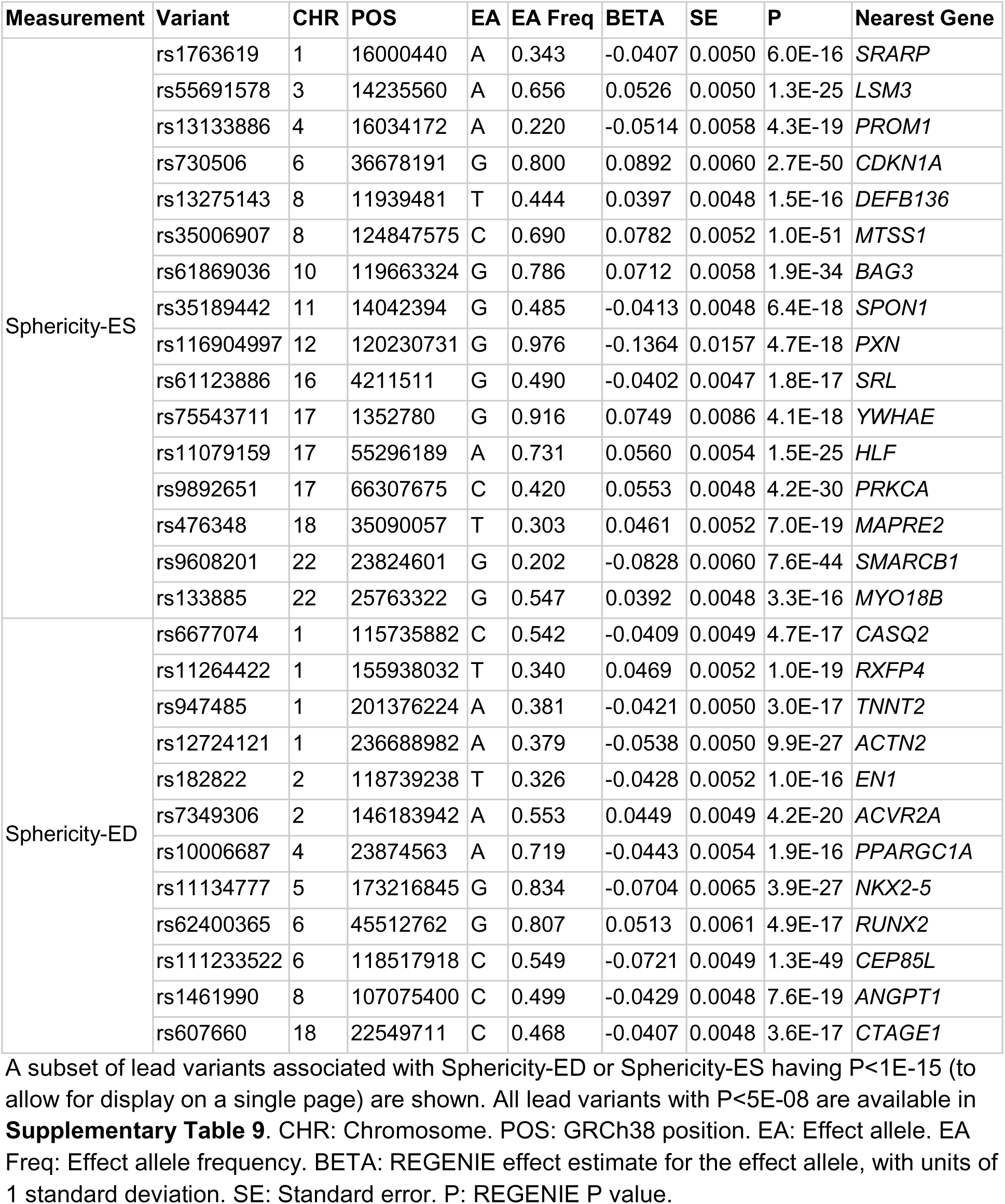
Loci associated with sphericity having P<5E-15.

| Measurement | Variant | CHR | POS | EA | EA Freq | BETA | SE | P | Nearest Gene |
| --- | --- | --- | --- | --- | --- | --- | --- | --- | --- |
| Sphericity-ES | rs1763619 | 1 | 16000440 | A | 0.343 | -0.0407 | 0.0050 | 6.0E-16 | *SRARP* |
|  | rs55691578 | 3 | 14235560 | A | 0.656 | 0.0526 | 0.0050 | 1.3E-25 | *LSM3* |
|  | rs13133886 | 4 | 16034172 | A | 0.220 | -0.0514 | 0.0058 | 4.3E-19 | *PROM1* |
|  | rs730506 | 6 | 36678191 | G | 0.800 | 0.0892 | 0.0060 | 2.7E-50 | *CDKN1A* |
|  | rs13275143 | 8 | 11939481 | T | 0.444 | 0.0397 | 0.0048 | 1.5E-16 | *DEFB136* |
|  | rs35006907 | 8 | 124847575 | C | 0.690 | 0.0782 | 0.0052 | 1.0E-51 | *MTSS1* |
|  | rs61869036 | 10 | 119663324 | G | 0.786 | 0.0712 | 0.0058 | 1.9E-34 | *BAG3* |
|  | rs35189442 | 11 | 14042394 | G | 0.485 | -0.0413 | 0.0048 | 6.4E-18 | *SPON1* |
|  | rs116904997 | 12 | 120230731 | G | 0.976 | -0.1364 | 0.0157 | 4.7E-18 | *PXN* |
|  | rs61123886 | 16 | 4211511 | G | 0.490 | -0.0402 | 0.0047 | 1.8E-17 | *SRL* |
|  | rs75543711 | 17 | 1352780 | G | 0.916 | 0.0749 | 0.0086 | 4.1E-18 | *YWHAE* |
|  | rs11079159 | 17 | 55296189 | A | 0.731 | 0.0560 | 0.0054 | 1.5E-25 | *HLF* |
|  | rs9892651 | 17 | 66307675 | C | 0.420 | 0.0553 | 0.0048 | 4.2E-30 | *PRKCA* |
|  | rs476348 | 18 | 35090057 | T | 0.303 | 0.0461 | 0.0052 | 7.0E-19 | *MAPRE2* |
|  | rs9608201 | 22 | 23824601 | G | 0.202 | -0.0828 | 0.0060 | 7.6E-44 | *SMARCB1* |
|  | rs133885 | 22 | 25763322 | G | 0.547 | 0.0392 | 0.0048 | 3.3E-16 | *MYO18B* |
| Sphericity-ED | rs6677074 | 1 | 115735882 | C | 0.542 | -0.0409 | 0.0049 | 4.7E-17 | *CASQ2* |
|  | rs11264422 | 1 | 155938032 | T | 0.340 | 0.0469 | 0.0052 | 1.0E-19 | *RXFP4* |
|  | rs947485 | 1 | 201376224 | A | 0.381 | -0.0421 | 0.0050 | 3.0E-17 | *TNNT2* |
|  | rs12724121 | 1 | 236688982 | A | 0.379 | -0.0538 | 0.0050 | 9.9E-27 | *ACTN2* |
|  | rs182822 | 2 | 118739238 | T | 0.326 | -0.0428 | 0.0052 | 1.0E-16 | *EN1* |
|  | rs7349306 | 2 | 146183942 | A | 0.553 | 0.0449 | 0.0049 | 4.2E-20 | *ACVR2A* |
|  | rs10006687 | 4 | 23874563 | A | 0.719 | -0.0443 | 0.0054 | 1.9E-16 | *PPARGC1A* |
|  | rs11134777 | 5 | 173216845 | G | 0.834 | -0.0704 | 0.0065 | 3.9E-27 | *NKX2-5* |
|  | rs62400365 | 6 | 45512762 | G | 0.807 | 0.0513 | 0.0061 | 4.9E-17 | *RUNX2* |
|  | rs111233522 | 6 | 118517918 | C | 0.549 | -0.0721 | 0.0049 | 1.3E-49 | *CEP85L* |
|  | rs1461990 | 8 | 107075400 | C | 0.499 | -0.0429 | 0.0048 | 7.6E-19 | *ANGPT1* |
|  | rs607660 | 18 | 22549711 | C | 0.468 | -0.0407 | 0.0048 | 3.6E-17 | *CTAGE1* |

**Table 3:** Low-frequency single-nucleotide variants.

| Variant | CHR | POS | EA | EA Freq | BETA | SE | P | Nearest Gene | Traits |
| --- | --- | --- | --- | --- | --- | --- | --- | --- | --- |
| rs188308364 | 2 | 32403593 | T | 0.9921 | -0.148 | 0.027 | 4.4e-08 | *BIRC6* | LVEF, Sphericity-ES |
| rs201672146 | 7 | 128854868 | G | 0.9966 | 0.243 | 0.041 | 2.4E-09 | *FLNC* | LVESV, LVEF, Surface Area-ES |
| rs112892337 | 8 | 134602310 | G | 0.9956 | -0.196 | 0.036 | 4.8E-08 | *ZFAT* | LVEDV |
| rs73653638 | 9 | 94683864 | T | 0.9978 | 0.313 | 0.056 | 1.7E-08 | *AOPEP* | Sphericity-ES |
| rs189569984 | 10 | 110784367 | C | 0.9915 | 0.177 | 0.026 | 9.6E-12 | *RBM20* | LVEDV, LVESV, LVEF, LV Mass, Surface Area-ED, Surface Area-ES |
| rs45550635 | 11 | 19192439 | A | 0.9952 | 0.190 | 0.034 | 2.2E-08 | *CSRP3* | LVESV, LVEF, Sphericity-ES |
| rs7308460 | 12 | 69413512 | C | 0.9989 | -0.443 | 0.075 | 3.6E-09 | *YEATS4* | LV Mass |
| rs533142647 | 20 | 12582994 | A | 0.9988 | -0.399 | 0.068 | 3.7E-09 | *SPTLC3* | LVESV, Surface Area-ES |
| rs545500184 | 20 | 33555658 | C | 0.9980 | 0.303 | 0.055 | 2.9E-08 | *CBFA2T2* | LV Mass |
| rs192896387 | 22 | 31093448 | A | 0.9960 | -0.223 | 0.037 | 2.2E-09 | *SMTN* | LVESV, Surface Area-ES |

Of the nine ACMG SF3.2 DCM genes^21^, eight were within 500kb of a GWAS locus (including *BAG3*, *DSP*, *FLNC*, *LMNA*, *RBM20*, *TNNC1*, *TNNT2*, and *TTN*, but not *DES*), as were three of the nine hypertrophic cardiomyopathy genes (sarcomeric genes *MYBPC3*, *MYH7*, and *TNNT2*).

### Sphericity common variant genetics

Aggregating the sphericity GWAS results at the gene level using MAGMA^22,23^, enrichment was identified for gene ontology terms related to heart and muscle development and sarcomere/I-band organization (**Supplementary Table 10**). Tissue-specific expression analysis using data from GTEx v10 showed the strongest associations with cardiac and arterial tissues (**Supplementary Figure 10**)^24,25^. Single-nucleus sequencing data from diseased human hearts^26^ showed significant association with cardiomyocytes and vascular smooth muscle cells (**Supplementary Figure 11**). Single-nucleus sequencing in normal hearts showed a pattern of cell-type enrichment in ventricular cardiomyocytes, atrial cardiomyocytes, and adipocytes for sphericity-ES and LVEF; in contrast, sphericity-ED was not associated with adipocytes but was instead enriched for genes expressed in fibroblast populations (**Supplementary Figure 12**)^27^.

With respect to individual loci, at the *E2F6* locus, the rs3922995-A allele was notable for being linked to greater sphericity in the present work (both -ES and -ED), to greater *E2F6* expression in the LV in GTEx^24^, and to greater risk for DCM in the DCM_Broad_ GWAS^9^. These findings are consistent with the observational relationship linking greater sphericity to greater DCM risk, as well as with animal models linking overexpression of *E2F6* to LV dilation and dysfunction^28^.

Another variant linked to sphericity-ED was rs947485 near *TNNT2*, which encodes cardiac troponin T. The reference A allele (MAF 0.38) was associated with lower sphericity-ED; the same allele forms a splicing quantitative trait locus (sQTL) associated with increased inclusion (reduced skipping) of exons 4 and 5 in atrial appendage tissue in GTEx v10^24,25^. Exon 5 of *TNNT2* is ordinarily spliced out of the adult heart^29^, and its inclusion in the fetal cardiac *TNNT2* transcript increases myofilament calcium sensitivity ^30,31^, suggesting a link between enhanced calcium sensitivity and reduced sphericity. Missense variants in *TNNT2* have also been associated with hypertrophic cardiomyopathy^32^ and enhanced calcium sensitivity^33^.

Highlighting the link between calcium handling and LV sphericity, additional genes involved in calcium handling, beyond *TNNT2*, were found at sphericity loci, including *CASQ2* (encoding calsequestrin-2, a sarcoplasmic reticulum calcium buffer and sensor), *SRL* (encoding sarcalumenin, another sarcoplasmic reticulum calcium buffer), *MICU2* (encoding ‘mitochondrial calcium uptake 2’ on the inner mitochondrial membrane), *SLC8A1* (encoding ‘sodium/calcium exchanger 1’ in the sarcolemma), and *STRN* (encoding striatin, which binds calmodulin).

Additional nearest-genes at sphericity loci likely play a more distant role in calcium modulation (e.g., *ALPK3*, *LDB3*, and others). Given the energetic demands of calcium regulation, the identification of variants near genes linked to mitochondrial energetics (*ECI2*, *SUGCT*, *RMDN2*, *TOP3A*, *PPARGC1A*, and the aforementioned *MICU2*) and metabolism (*BCAT1*, *ASPG*, *SLC38A2*, *ADK*, and *TKT*) was also notable.

### Contextualization with previous DCM and LV measurement GWAS

A recent large-scale GWAS (14,256 DCM cases and 1,199,156 controls) identified 26 loci associated with DCM (“DCM_Broad_”) and 12 loci within more strictly defined DCM cases (“DCM_Narrow_”, **Supplemental Table 11**)^9^. For DCM_Narrow_, 11 of the 12 loci were within 500kb of a variant linked to LV structure and function in the present study with P<5E-08; only the locus near *PHB1* (MAF 0.018) was not. For DCM_Broad_, 22 of the 26 loci were within 500kb of variants associated at P<5E-08 for at least one LV trait in the present study—and for 20, the specific DCM_Broad_ lead variant had P<5E-08 in the present study. The LV traits sharing the most loci with DCM_Broad_ were LVEF, with 21, and sphericity-ES, with 19. The four DCM_Broad_ loci without nearby variants having P<5E-08 in the present study were those near *AXDND1*, *PDLIM5*, *PITX2*, and *CAMK2D*. Of these, the *CAMK2D* and *PDLIM5* loci had variants with P<1E-04 for at least one LV trait; the *PDLIM5* and *AXDND1* variants had low MAF (0.017-0.018); and the *PITX2* variant was more strongly associated with atrial fibrillation than with DCM_Broad_ (**Supplementary Table 12**)^34^.

Genetic associations were also reassessed from a 2020 GWAS for LV structure and function, which consisted of a subset of 36,000 participants from the present study^8^. Study-wide, 43 of the 46 loci associated with LVEDV, LVESV, or LVEF from the 2020 analysis remained significant with P<5E-08 in the present study; the three that did not were near *HECTD4*, *ATP5SL*, and *RRAS2*. Sub-significant loci were also reassessed: 16 of 23 LVEF loci with P values between 1E-06 and 5E-08 newly achieved P<5E-08 in the present study, as did 25 of 38 for LVEDV and 24 of 31 for LVESV.

### Olink proteomic Mendelian randomization

A *cis*-proteomic Mendelian randomization was conducted using variants within 944 kilobases^35^ of the genes encoding circulating proteins from the Olink Explore 3072 protein panel^36^ as the exposure instruments and the LV GWAS as outcome instruments. In the primary analysis using the Wald ratio (when only one independent variant was available) or inverse variance weighting (IVW), 279 protein-trait pairs (121 distinct proteins) were significantly associated with at least one LV measurement at a study-wide false discovery rate (FDR) of 0.05 (**Supplementary Figure 13**, **Supplementary Table 13**). 33 proteins were associated with sphericity-ES and 22 with sphericity-ED (45 distinct proteins).

The strongest statistical association was between genetically mediated levels of ZBTB17 and LVEF (-1.1 SD per SD, P=3.9E-28). However, the Steiger test^37^ did not support a causal association. Notably, in GTEx v10^24,25^, the genetic variant driving this result (rs848217) is also a multi-tissue eQTL for *HSPB7* and *CLCNKA*. Neither protein is measured on the Olink panel; therefore the association may be driven by horizontal pleiotropy through the unmeasured proteins. No other significantly associated proteins failed the Steiger test. The next strongest associations with LVEF by effect size were MSRA and BACH1: for both, greater genetic dosage of the protein was associated with genetic reduction in LVEF. The MSRA association was driven by one variant (rs12546887) which lies in a complex region of the genome near *GATA4* and forms eQTLs for dozens of genes whose protein products are not included on the Olink panel.

The BACH1 association was driven by rs368322, intronic to the *BACH1* gene and an eQTL for *BACH1* in fibroblasts and cardiac tissue. Prior work has linked deletion of *bach1* in mice to LV protection during pressure overload^38^, and degradation of the BACH1 protein with hemin to increased peri-infarct revascularization in a mouse model^39^.

21 of the 121 associated proteins mapped to known drug targets from the OpenTargets platform^40^: AGER, CA9, CD27, DPEP1, ENG, FCGR2B, FGFR4, FN1, FOLH1, GAS6, HSPA1A, IL12B, IL6R, NECTIN4, NOS3, PARP1, RSPO3, SOST, TFPI, TNFSF12, and TP53. For example, greater genetically mediated PARP1 was associated with lower sphericity-ES and sphericity-ED, and greater LVEF (all directionally concordant with greater risk for hypertrophic cardiomyopathy). Directionally consistent with this observation, prior literature has linked inhibition of PARP1 to reduced risk for cardiac hypertrophy in animal models of cardiac hypertrophy^41,42^. PARP1 inhibitors are in clinical use as antineoplastic agents, such as olaparib^43^.

A prior large-scale effort linked LV structure and function to seven proteins, including SPON1, IGFBP7, CCL15, FSTL3, ANGPTL3, NRP1, and MFAP4^44^. All seven proteins were tested in the present analysis, but only the SPON1 association was affirmatively validated. In the present study, genetically mediated greater circulating levels of SPON1 were associated with greater LVEF (P=3.8E-19) and reduced LV volume, surface area, and sphericity.

### Genes harboring rare genetic variation for LV structure and function

Rare (MAF < 0.001) coding variants, including LOFTEE high-confidence loss-of-function (LoF) variants and missense variants predicted to be deleterious by AlphaMissense^45,46^, were aggregated at the gene level for burden testing (**Methods**). 17 genes were associated with at least one LV measurement: *ALPK3*, *CDH1*, *CEACAM4*, *CEP78, CSRP3*, *DPY19L4*, *EHD2*, *FLNC*, *MYBPC3*, *NOS3*, *PKP2*, *RNF40*, *RRAD*, *SYNM*, *TNNT2*, *TTN*, and *ZNF189* (**Figure 5**).

**Figure 5:**
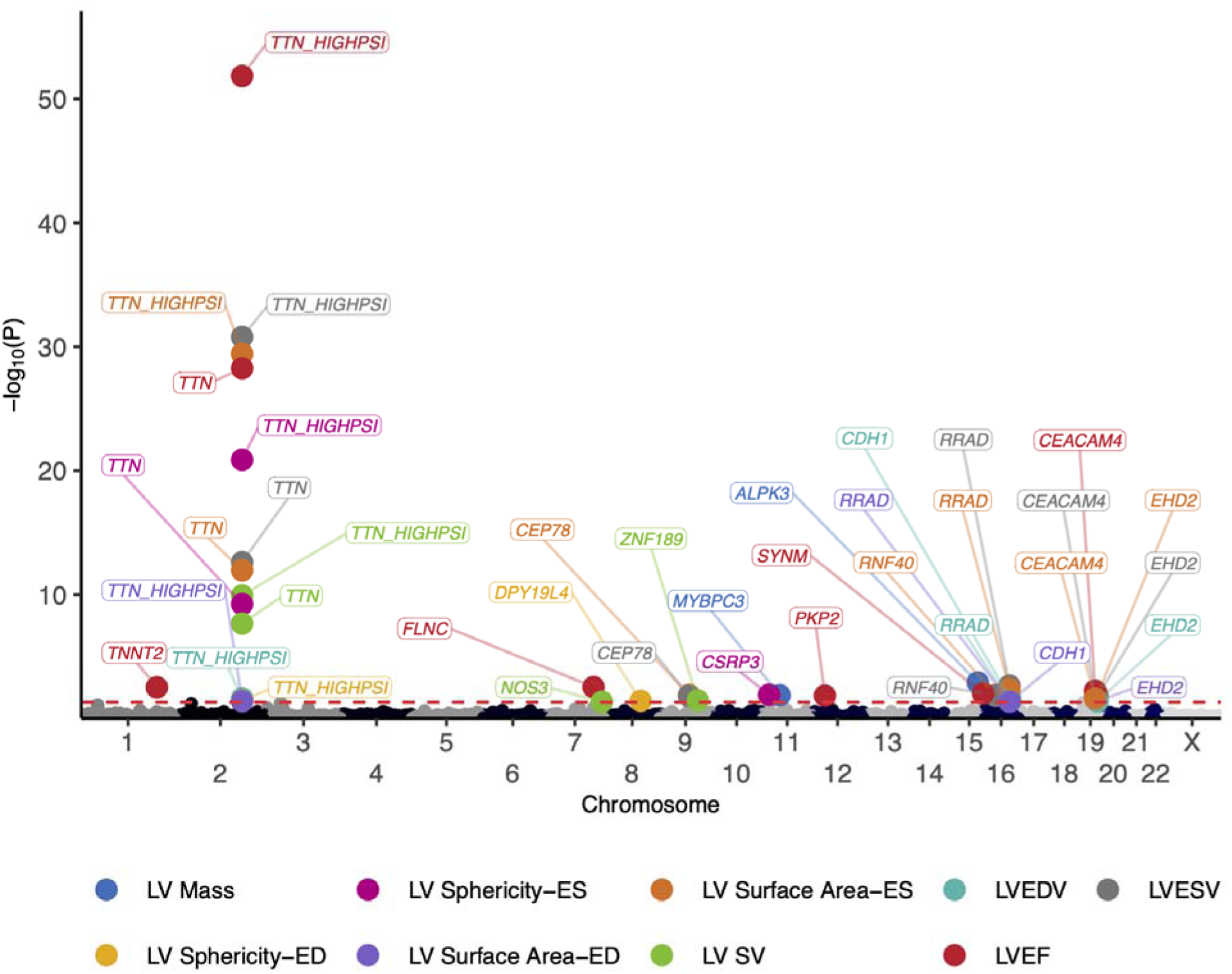
Rare variant associations. Chromosomal position (x-axis) and -log10 gene-level Benjamini-Hochberg-adjusted P value (y-axis) are depicted. Note that *TTN_HIGHPSI* represents a subset of the *TTN* transcript that includes only exons that are highly present cardiac mRNA transcripts^47^.

All LV measurements except for LV mass were associated with LoF variants in *TTN*, particularly in the exons that are highly spliced into cardiac transcripts (TTNtv)^47^. The strongest statistical association was between LVEF and TTNtv (-0.84 SD of LVEF among carriers; P=1.6E-56), consistent with many prior reports^8,9,48,49^. In addition to TTNtv, LVEF was associated with variants in two additional ACMG SF v3.2^21^ DCM-linked genes: *FLNC* and *TNNT2*, as well as the SF v3.2 arrhythmogenic right ventricular dysplasia-linked *PKP2*. LVEF was also linked to rare variant burden in *SYNM*, encoding the intermediate filament synemin protein that interacts with desmin and has been linked to DCM in humans and LV dysfunction in mice^50,51^.

In addition to *TTN*, sphericity-ED was linked to rare variant burden in *DPY19L4*. Sphericity-ES was linked to variants in *TTN* and *CSRP3*, which encodes a cardiac LIM protein that has been associated with hypertrophic cardiomyopathy^52^.

LV mass was associated with rare variants in the ACMG SF v3.2 HCM gene *MYBPC3*, as well as *ALPK3*, which has also been linked to HCM^53^. Both LVEDV and LVESV were associated with variants in *EHD2*, whose protein product regulates ATP-sensitive potassium channel expression in the sarcolemma^54^, and with variants in *RRAD*, whose protein product regulates calcium signaling^55^. Variants in *RRAD* have previously been linked to Brugada syndrome^56^. Additional associations are listed in **Supplementary Table 14** and **Figure 5**.

To assess disease relevance, in the 406,754 UK Biobank participants who did not undergo imaging, variants in the 17 genes associated with LV structure and function were tested for association with DCM, HCM, and heart failure. Nominal (unadjusted P<0.05) evidence was found between at least one disease and *TTN*, *MYBPC3*, *ALPK3*, *TNNT2*, *FLNC*, *RRAD*, *CEACAM4*, *PKP2*, *CEP78*, *NOS3*, and *RNF40* (**Supplementary Table 15**). The largest effect sizes were observed for LoF variation in *MYBPC3* for HCM (635 events, 208 variant carriers, 30 events in carriers, odds ratio [OR] 118, P=2.0E-121) and TTNtv for DCM (980 events, 1,670 variant carriers, 90 events in variant carriers, OR 26.4, P=1.2E-180).

External validation was also pursued in 410,193 *All of Us* participants. Nominal (unadjusted P<0.05) evidence was found for *MYBPC3*, *TTN*, *FLNC*, *TNNT2*, *RRAD*, *ZNF189*, *CEACAM4*, *CSRP3*, *PKP2*, and *ALPK3*, with the largest effect sizes again seen for *MYBPC3* (OR 49.1 for HCM) and TTNtv (OR 13.4 for DCM) (**Supplementary Table 16**). For *All of Us* participants with TTNtv, the estimated cumulative incidence of DCM by age 80 was 33% (**Supplementary Figure 14**).

### Polygenic scores predict DCM and HCM

Genome-wide polygenic scores for each of the LV measurements were constructed from the 1.4 million HapMap3+ variants^57^ using PRScs^58^ (**Methods**) and then tested for disease association in up to 406,754 UK Biobank participants without imaging (**Supplementary Table 17**) and 410,193 *All of Us* participants.

The LVESV score was most strongly associated with DCM (781 cases; HR 1.70 per SD, 95% CI 1.58-1.82, P=9.2E-49; **Figure 6**). Compared to all other individuals in the non-imaging UK Biobank cohort, those in the top 5% of the LVESV score had an HR of 3.3 (95% CI 2.7-4.0, P=1.1E-31). In external validation in *All of Us*, the LVESV score had an HR of 1.53 per SD for DCM in EUR-ancestry individuals (P=1.4E-14, 330 events and 230,531 non-events), which remained comparable when all individuals were tested jointly (HR=1.48 per SD, P=5.0E-24, 684 events and 409,509 non-events).

**Figure 6:**
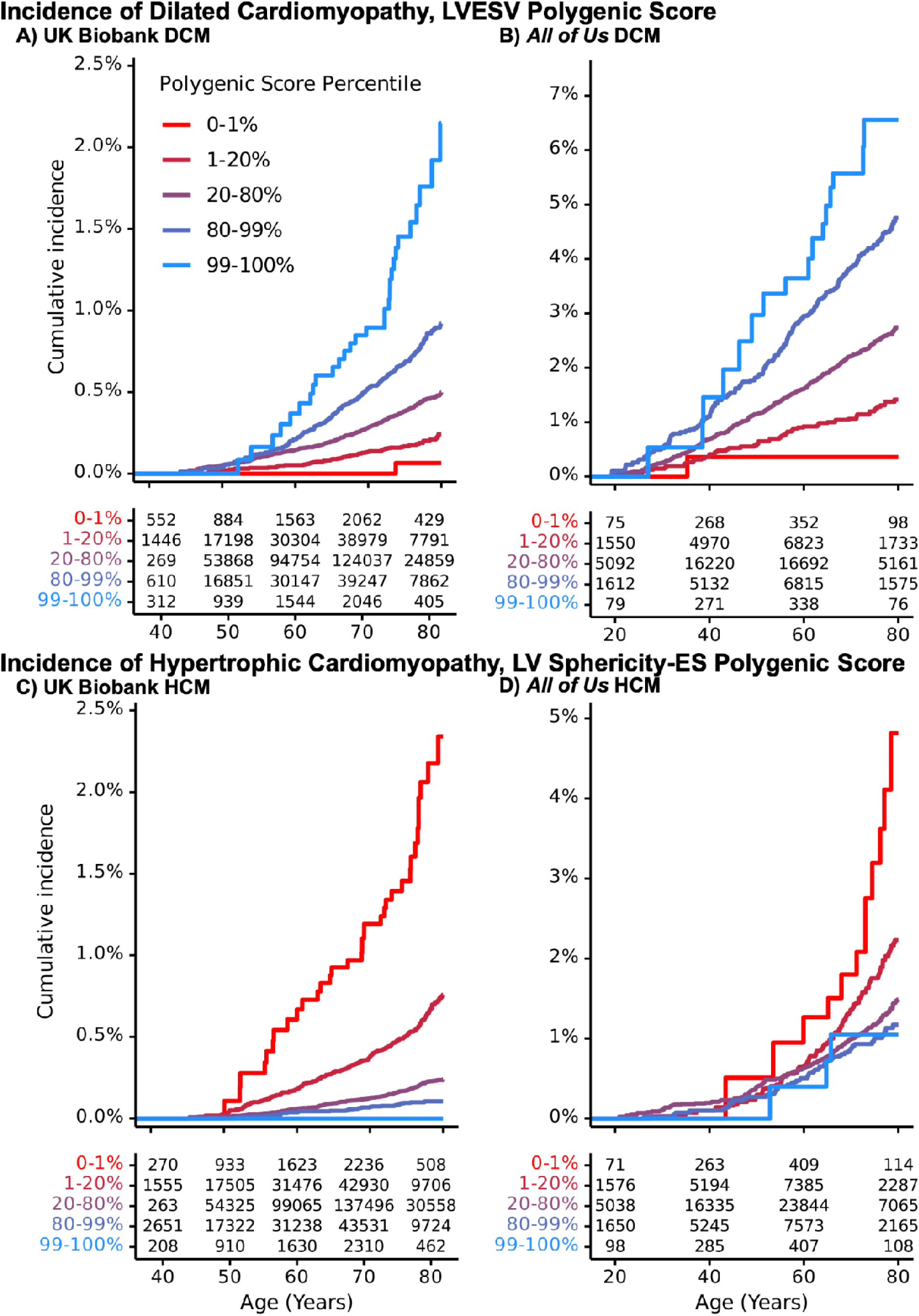
Polygenic risk for dilated & hypertrophic cardiomyopathy. Age-specific cumulative incidence by polygenic score quantile. Curves are left-truncated at each participant’s enrollment age, and therefore depict risk conditional on being alive, enrolled, disease-free, and uncensored at that attained age. **Panels**: Incident dilated cardiomyopathy (DCM) by quartiles of the LVESV polygenic score in UK Biobank participants without imaging (**Panel A**) and *All of Us* (**Panel B**). Incident hypertrophic cardiomyopathy (HCM) by quartiles of the LV sphericity-ES polygenic score in UK Biobank participants without imaging (**Panel C**) and *All of Us* (**Panel D**).

For HCM, a 1-SD increase in the sphericity-ES score was associated with a lower risk of incident disease (530 cases; HR 0.47 per SD, 95% CI 0.43-0.51, P=5.6E-71). Compared to the remainder, those in the bottom 5% of the sphericity-ES score had an HR of 4.4 for HCM (95% CI 3.5-5.4, P=3.7E-39). In *All of Us*, the sphericity-ES score had an HR of 0.64 per SD for HCM in EUR-ancestry individuals (P=9.7E-14, 272 events and 230,589 non-events), or 0.72 per SD in all individuals (P=2.9E-12, 448 events and 409,745 non-events).

To contextualize these imaging-derived polygenic scores with recently developed genome-wide polygenic risk scores (PRS) for DCM^9^ and HCM^59^, all scores were applied to the same 406,754 UK Biobank participants without imaging and tested for association with DCM and HCM. The LV sphericity-ES score (HR 1.57 per SD, 95% CI 1.46-1.69) was as strongly associated with DCM as the DCM PRS (HR 1.55 per SD, 95% CI 1.43-1.67) by the non-nested Cox likelihood ratio test (two-tailed P=0.18 for unequal fit)^60^. Notably, the LVESV score described above (HR 1.70 per SD) was superior (two-tailed P=1.7E-03) to the DCM PRS (HR 1.55 per SD). For HCM, the LV sphericity-ES score (HR 2.15 per SD decrease, 95% CI 1.98-2.34) was as strongly associated (two-tailed P=0.72) as the HCM PRS (HR 2.19 per SD, 95% CI 2.02-2.39). When benchmarked against multi-trait analysis of GWAS (MTAG)–enhanced PRS from the same DCM and HCM studies, the LVESV score was non-inferior to the MTAG-DCM PRS for DCM (two-tailed P=0.80), and the LV sphericity-ES score was non-inferior to the MTAG-HCM PRS for HCM (two-tailed P=0.13).

## Discussion

This work assessed 3D LV sphericity—a measurement that formalizes how closely the LV blood pool approximates a sphere—and found it to be an independent and heritable risk marker for dilated cardiomyopathy and hypertrophic cardiomyopathy. Assessing LV sphericity alongside the traditional LV measurements enabled the identification of 366 distinct common genetic loci (1,115 locus-trait pairs) using whole genome sequencing in 80,389 UK Biobank participants.

The high degree of locus overlap with a recent large-scale DCM GWAS of over 1 million participants raises the possibility that most of the loci that will eventually be discovered in future DCM GWAS are already available in the present analyses of LV structure and function.

The approach to measuring sphericity used in this work, based on a 3D mesh representation of the LV blood pool, took advantage of a simple formula—the surface area of an equal-volume sphere divided by the true LV surface area^18^—in both diastole and systole. These features distinguish it from prior work in end-diastole that evaluated the 2D aspect ratio of the LV from the 4-chamber view (width vs height)^6^, the long-axis length and volume of the LV^4,5^, or surface mesh principal components (qualitatively interpreted as being related to sphericity)^61^. In the present work, sphericity-ES yielded a greater effect estimate for incident DCM than prior sphericity measurements—or even standard LV measurements such as LVEF. One possible explanation for the sphericity-DCM link being stronger than the LVEF-DCM link could be that remodeling towards a more globular shape begins earlier than the decrement in LVEF; while this cannot be definitively assessed in this cross-sectional study, it is an avenue for future inquiry that has clinical implications. From a technical perspective, LV sphericity-ES has the appealing characteristics of being dimensionless, and only requiring measurement at one moment; this latter point distinguishes sphericity-ES from LVEF, which—though also dimensionless—requires measurement both at end-systole and end-diastole, compounding measurement error from both.

This approach to measuring 3D sphericity also dramatically improved power for genetic discovery (100 genetic loci for LV sphericity-ES in ∼80,000 participants vs four loci for the LV aspect ratio in ∼36,000 participants). The common variant analyses in the present work using WGS data in 80,389 individuals confirmed nearly every previously reported LV locus with P<5E- 08 from earlier releases of UK Biobank imaging, and substantially expanded the number of distinct loci associated with LV structure and function to 366, without depending on multitrait statistical assumptions. They also permitted an exploration of sub-significant loci with P between 1E-06 and 5E-08 from a prior 36,000-participant LV GWAS^8^: more than two-thirds of these loci strengthened to achieve P<5E-08 in the present study. This suggests that, despite elevated noise above the P=5E-08 threshold, there is also a substantial amount of information in that range, and it is reasonable to consider such evidence when selecting loci for functional validation.

In addition to recovering most of the loci identified in a recent large-scale DCM GWAS^9^ (10 of 11 DCM_Narrow_ loci and 22 of 26 DCM_Broad_ loci), the common variant analyses in the present work also yielded a polygenic score (for LVESV) that was more strongly predictive of DCM than the DCM-specific polygenic score, and was non-inferior for predicting DCM compared to the MTAG-enhanced DCM polygenic score. Notably, one score (the sphericity-ES score), was equally informative for DCM as the DCM score^9^ and equally informative for HCM as the HCM score^59^ when tested in the same participants. This underscores the opposing relationship for DCM and HCM at the extremes of LV sphericity, both for the measured phenotype and for its polygenic score. These findings also emphasize the sample efficiency of quantitative traits for studying the genetic basis for human disease: genetic analyses of quantitative LV measurements in 80,000 people yielded similarly powerful instruments as those derived from over 1 million individuals with binary traits. For those at the extremes of polygenic risk, the cumulative incidence of DCM by age 80 was one fifth as high as that of TTNtv carriers (6.6% cumulative incidence for the top 1% of the LVESV score vs 33% for TTNtv carriers; **Supplementary Figure 14**). While rare variant burden has an outsized impact on a small number of individuals, polygenic risk can quantitatively contribute to comprehensive risk assessments for all individuals. Consideration of both common and rare variation is warranted in future cardiomyopathy risk scores, particularly for young individuals with few observable risk factors.

The use of WGS in this study facilitated the evaluation of low-frequency variants (MAF 0.001-0.01) in the GWAS. Several low-frequency missense variants were identified in known cardiomyopathy-related genes (*RBM20*, *CSRP3*, and *FLNC*) with effect sizes of approximately 0.2-0.25 SD of the underlying LV measurement. These are larger than the effect sizes of the common variants in the study (mean effect estimate 0.04 SD; **Figure 4**), but smaller than those of rare variants (up to 1.1 SD; **Supplementary Table 14**). While several of the variants in the low frequency range have previously been evaluated in clinical genetics laboratories and considered benign (e.g., *RBM20* p.Ser455Leu), the observations in the present work suggest that they effectively function in a quantitative role that is similar to that of common variants, but with 5-fold greater effect. Efforts to comprehensively incorporate low frequency variants into polygenic scores, particularly given the potential limitations from ancestry-specific allele frequency differences, will be of interest in the future.

Of the 17 genes associated with at least one LV measurement in the rare variant burden test, 11 were within 500kb of a GWAS lead variant (*ALPK3*, *CSRP3*, *DPY19L4*, *EHD2*, *FLNC*, *MYBPC3*, *NOS3*, *RRAD*, *SYNM*, *TNNT2*, and *TTN*), and six of these were also the nearest gene to the lead variant at the locus for the same trait (*ALPK3*, *CSRP3*, *FLNC*, *MYBPC3*, *NOS3*, and *TTN*). At the four loci where the gene harboring a rare variant signal was not the nearest gene to the lead common variant, different traits were flagged by the common- and rare-variant analyses. For example, *SYNM* harbored rare variants associated with LVEF, whereas the common variant signal within 500kb had a lead variant that was intronic to *IGF1R* and was associated with LV mass, which fits with prior evidence suggesting that increased *IGF1R* signaling in cardiomyocytes increases LV mass^62^. While most of the associated genes had extensive prior support for cardiac relevance, some findings do provide consilient support for genes with modest prior evidence. For example, *DPY19L4*, associated here with sphericity- ED, was recently identified as the nearest gene to the lead common variant from a heart failure GWAS in Japanese participants^63^. Nevertheless, even with quantitative traits in 80,000 participants, the power to make novel rare variant-based discoveries remains limited; therefore, in the rare-variant regime in particular, large-scale cardiomyopathy sequencing analyses are of considerable future interest.

One note of caution is warranted based on the proteomic Mendelian randomization analyses. While genetically mediated levels of 121 distinct proteins were putatively causally associated with LV structure and function, several of the strongest associations had evidence for pleiotropy, including the single strongest association (ZBTB17, which failed the Steiger test). Because not every protein is assayed in the current proteomic panels, particular caution is warranted when interpreting these analyses, and triangulation with other data, such as expression from GTEx, is warranted. Nevertheless, the proteomic findings provide supportive evidence to previously reported proteins such as RSPO3^64^ and SPON1^65^, and nominate dozens of other proteins for further investigation.

In conclusion, 3D LV sphericity is a novel measurement that has strong epidemiologic and polygenic associations with HCM and DCM. Using WGS in 80,000 individuals, LV structure and function were associated with 366 loci with common- and low-frequency variants, and with 17 genes harboring rare genetic variants.

### Limitations

While the SAX deep learning model was externally validated in ACDC^15^, the long-axis deep learning models have not been externally validated and may not generalize beyond the MRI scanners and imaging protocols used in the UK Biobank. Future work will be required to validate the epidemiological associations between sphericity and disease in other cohorts, especially with more commonly used clinical imaging modalities such as echocardiography. The proteomic Mendelian randomization will be biased towards the observational associations due to partial sample overlap between the Olink GWAS and the MRI GWAS. The rare variant analyses were reported with false discovery control within each phenotype, rather than across all tests. The Kaplan-Meier cumulative incidence estimates in *All of Us* spanning ∼60 years of age nevertheless represent less than 4 years of follow-up time for each participant, on average, and should therefore be viewed as broad estimates. UK Biobank participants predominantly have genetic identities similar to individuals from Europe, which limits generalization to individuals with other genetic identities, particularly with respect to polygenic scores.

## Methods

### Study design

Deep learning model development and epidemiologic and genetic analyses were conducted in the UK Biobank. External validation was conducted in *All of Us*. Study protocols complied with the tenets of the Declaration of Helsinki. All UK Biobank participants provided written informed consent^66^, and only those participants who had not withdrawn consent as of October 2023 were analyzed. UK Biobank analyses were conducted under application #41664. Each *All of Us* biobank participant provided written informed consent. The analyses were approved by the UCSF Institutional Review Board, #22-37715.

### Statistical analyses

Analyses were conducted in R version 4.3.3 (R Foundation for Statistical Computing, Vienna, Austria) unless otherwise stated. Disease status was defined based on diagnostic and procedural codes (**Supplementary Table 18**). Prevalent disease analyses treated each heart measurement as the dependent variable in a standard linear model with disease status as the independent variable of interest, with covariate adjustment for age at the time of imaging, sex, and the imaging center using *lm* in R. Incident disease analysis was conducted using Cox proportional hazards models using the R *survival* package. Individuals with events or exclusion criteria occurring prior to MRI were excluded from the analysis. The date of MRI was treated as the beginning of follow-up time, which terminated at the event occurrence or was censored at the first occurrence of any of the following: an exclusion criterion; loss to follow-up; death; or the centrally recommended censoring date from the UK Biobank. For mortality in heart failure, the same Cox modeling approach and covariates were used, with an additional interaction term for the presence of heart failure at the time of imaging and the sphericity-ES values, with mortality as the outcome. For polygenic score-based analyses in participants without imaging (UK Biobank and *All of Us*, separately), Cox models were built with follow-up time starting at enrollment, ending at the first event occurrence or censoring, with the scaled polygenic score as the independent variable of interest and covariates including the first 10 principal components of ancestry, age, and sex.

### Study population and magnetic resonance imaging

The UK Biobank is a prospective cohort study that enrolled over 500,000 individuals in the UK aged 40-69 years between 2006-2010^10^. Comprehensive phenotyping data were obtained for each participant, and inpatient electronic health records and death registries are linked to the cohort^10,67^.

The imaging substudy of the UK Biobank used 1.5 Tesla Siemens MAGNETOM Aera magnetic resonance imaging (MRI) scanners (Siemens Healthineers, Erlangen, Germany)^11^. The cardiac cycle was represented by 50 evenly temporally spaced images for each view. The three long-axis imaging views (2-, 3-, and 4-chamber) each represented a distinct plane. The short-axis images represented an evenly spaced volumetric stack of images along the long axis of the left ventricle; the number of planes in this stack was chosen at the time of imaging and dependent on the size of the heart. All data were represented in the DICOM format which contained sufficient metadata to register each pixel into a co-oriented 3D space (**Supplementary Methods**).

### Deep learning to label LV pixels from MRI

Four distinct multiscale vision transformer-based semantic segmentation (pixel-labeling) deep learning models were built in PyTorch^68–70^ and trained using data manually labeled by a cardiologist (JPP): one each for the two-, three-, and four-chamber long axis MRI planes and a fourth for the short axis MRI stack. The hyperparameters and model training are detailed in the **Supplementary Methods**. With these models, the left atrial blood pool, the LV blood pool, and the LV myocardium were labeled in each image.

### LV surface reconstruction and measurements

3D reconstruction from the semantic segmentation output was performed using the screened Poisson surface reconstruction algorithm in a manner consistent with previous use of this algorithm for left atrial reconstruction^16,17,71^, yielding LV surface area and volume (**Supplementary Methods**). The reconstruction process was repeated at each of the 50 time points during the cardiac cycle for each individual, yielding LV blood pool surface area and volume at each step. The greatest volume was defined to be the LV end diastolic volume (LVEDV) and the point at which LV sphericity at end-diastole was measured; the smallest volume was defined to be the LV end systolic volume (LVESV) and the point at which LV sphericity at end-systole was measured. Stroke volume was defined as LVEDV minus LVESV. LV ejection fraction (LVEF) was defined as stroke volume divided by LVEDV. LV mass was measured using an analogous approach, as detailed in the **Supplementary Methods**.

### Definition of 3D LV sphericity index

To define the 3D LV sphericity index, the approach of Wadell was used: the sphericity index is the surface area of a sphere having the same volume as the LV blood pool, divided by the measured surface area of the LV blood pool^18^. For comparative purposes, LV sphericity was also computed using the length-volume method^4,5^. Formulas are presented in the **Supplementary Methods**.

### Quality control

Participants were excluded from analysis due to gating errors, transient motion artifacts, and geometric holes in the reconstructed blood pool (**Supplementary Methods**). For the GWAS, additional exclusions were made based on disease history and standard genetic QC (**Supplementary Methods**).

### UK Biobank whole genome sequencing overview

All genetic analyses were conducted with respect to GRCh38 using whole genome sequencing (WGS) data, which were available for 490,036 UK Biobank participants after central quality control^72^ (**Supplementary Methods**). All variants with a non-PASS filter were excluded before analysis in the present study.

### Single-variant genetic analyses

To create a suitable variant panel for the REGENIE Step 1 procedure, the variant sites from the original UK Biobank genotyping data^67^ (UK Biobank field ID #100315) were lifted over to GRCh38^73^, and the 706,467 available sites were then extracted from the WGS data. Each phenotype was residualized for age, age^2^, sex, and the first 10 principal components of ancestry, the MRI scanner, and the atrial motion category (**Supplementary Methods**). REGENIE v4.1^74^ was used to conduct single-variant analyses for each of the nine phenotypes for all variants with minor allele frequency (MAF) >= 0.001.

### Olink cis-proteomic Mendelian randomization

*Cis*-protein quantitative trait locus (*cis*-pQTL) data were generated from the Sun, *et al*, 2023 summary statistics derived from Olink proteomic measurements in 35,000 European-ancestry UK Biobank participants^36^. The full summary statistics were reprocessed to retain variants with P<5e-08 that were within 944kb of the target gene and therefore eligible to be considered “*cis*” ^35^. To obtain non-redundant variants, clumping was then performed with the WGS data from all UK Biobank participants using plink2 --clump with an r^2^ threshold of 0.01^75^. Two-sample Mendelian randomization was then performed in R 4.5.0 using *TwoSampleMR* v0.6.16, treating the proteins as the exposure and the LV measurements as the outcomes^76^. The inverse-variance weighted method (or Wald method, when only one variant was available) was treated as the primary analysis, with the other default methods treated as sensitivity analyses. Study-wide Benjamini-Hochberg FDR was applied across all protein-trait pairs^77^.

### Rare genetic variant burden testing

Variants were extracted from the WGS data within exons defined by MANE v1.4 tracks^78^, plus 10 base pairs on either side to allow for splice sites. Variants were annotated with VEP v110^79^, and those annotated as LOFTEE-HC or LOFTEE-LC loss-of-function variation, or AlphaMissense likely-pathogenic variation^45,46,80^, were retained. Burden masks were assembled using LOFTEE-HC alone, LOFTEE-LC and AlphaMissense, and all three together. Transcripts having fewer than 2 variant sites, or having fewer than 100 total variant allele carriers across all three annotation types, were excluded. An additional gene mask was generated for *TTN* variants within exons that are highly spliced into cardiac transcripts^47^, here termed *TTN_HIGHPSI*.

Burden and SKAT-O tests were implemented with REGENIE v4.1^74,81,82^. In both Stage 1 and Stage 2, nine polygenic scores—comprising the weighted allelic sum of the lead variants from the GWAS for each trait—were included in both Stage 1 and Stage 2 to improve power^83^. In addition, adjustment was made for height, weight, age, age^2^, sex, and the first 10 principal components of ancestry, the MRI scanner, and the atrial motion category (**Supplementary Methods**). Burden masks were generated for all variants with MAF<0.001 and for singleton variants. Because REGENIE produces a score per {phenotype, transcript, mask, frequency}, P-values were aggregated per gene-phenotype pair using ACAT^84^. The false discovery rate (FDR) was controlled per phenotype using Benjamini-Hochberg adjustment with R’s *p.adjust*^77^. Genes that were FDR-significant were tested for association with heart failure, DCM, and HCM in the UK Biobank participants who did not undergo imaging, adjusting for age and sex as covariates.

### Polygenic scores

For each LV measurement GWAS, genome-wide polygenic scores were computed with PRScs^58^ using a 1,408,687-million variant panel based on the expanded HapMap3+ variant list^57^ using *phi=’auto’* (**Supplementary Methods**). Scores were then PC-residualized in the 1000 Genomes participants^85–87^ and projected into UK Biobank participants.

PRS from the recent DCM (PGS004861) and HCM (PGS004910) GWAS, and the MTAG versions of the same (PGS004862 and PGS004911, respectively) were fetched from the PGSCatalog^88^ and applied in UK Biobank participants without imaging for comparison.

## Code availability

Trained PyTorch semantic segmentation deep learning models will be available at doi:10.5281/zenodo.16128460 upon final publication.

## Data availability

GWAS summary statistics will be available in the GWAS Catalog upon final publication. Polygenic score weights will be available in the PGS Catalog upon final publication. Individual-level data in the *All of Us* biobank and UK Biobank are accessible to approved researchers. Individual-level measurements from the present work will be returned to UK Biobank for use by other approved researchers following UK Biobank protocols. Linkage disequilibrium matrices for applying the HapMap3+ variant panel with PRScs will be available at doi:10.5281/zenodo.16132532 upon final publication.

## Data Availability

GWAS summary statistics will be available in the GWAS Catalog upon final publication. Polygenic score weights will be available in the PGS Catalog upon final publication. Individual-level data in the All of Us biobank and UK Biobank are accessible to approved researchers. Individual-level measurements from the present work will be returned to UK Biobank for use by other approved researchers following UK Biobank protocols. Linkage disequilibrium matrices for applying the HapMap3+ variant panel with PRScs will be available at doi:10.5281/zenodo.16132532 upon final publication.

## Acknowledgments

This work was supported by NIH R01HL178603, NIH K08HL159346, and by institutional support from the University of California San Francisco.

This research has been conducted using the UK Biobank Resource under application #41664. This work uses data provided by patients and collected by the NHS as part of their care and support. UK Biobank is generously supported by its founding funders the Wellcome Trust and UK Medical Research Council, as well as the British Heart Foundation, Cancer Research UK, Department of Health, Northwest Regional Development Agency and Scottish Government.

*All of Us* participants are gratefully acknowledged for their contributions; without them, this research would not have been possible. The National Institutes of Health’s *All of Us* Research Program is also gratefully acknowledged for making available the participant data examined in this study. The *All of Us* Research Program is supported by the National Institutes of Health, Office of the Director: Regional Medical Centers: 1 OT2 OD026549; 1 OT2 OD026554; 1 OT2 OD026557; 1 OT2 OD026556; 1 OT2 OD026550; 1 OT2 OD 026552; 1 OT2 OD026553; 1 OT2 OD026548; 1 OT2 OD026551; 1 OT2 OD026555; IAA #: AOD 16037; Federally Qualified Health Centers: HHSN 263201600085U; Data and Research Center: 5 U2C OD023196; Biobank: 1 U24 OD023121; The Participant Center: U24 OD023176; Participant Technology Systems Center: 1 U24 OD023163; Communications and Engagement: 3 OT2 OD023205; 3 OT2 OD023206; and Community Partners: 1 OT2 OD025277; 3 OT2 OD025315; 1 OT2 OD025337; 1 OT2 OD025276.

