## Supplementary Note for "Whole-genome sequencing analysis of left ventricular structure and sphericity in 80,000 people"

### Supplementary Methods

#### Definition of 3D left ventricular sphericity index

The approach of Wadell (1935) was used to define 3D left ventricular sphericity^1^. **Equation 1** produces an estimate of the surface area of a perfect sphere with a volume equivalent to that of the left ventricular (LV) blood pool. **Equation 2** defines sphericity index based on the sphere’s surface area (from **Equation 1**) and the left ventricular blood pool’s surface area (the measured quantity). This dimensionless measure of sphericity is bounded between a minimum value of 0 and a maximum value of 1.

**Equation 1 (surface area of sphere with equivalent volume)**

$$4\pi\times\left( \sqrt[3]{\frac{3 \times Volume}{4\pi}} \right)^{2}$$

**Equation 2 (Wadell’s sphericity index)**

$$\frac{Surface area of sphere with equivalent volume as the LV, from Equation 1}{LV surface area}$$

For comparison with prior work, a second measure of LV sphericity was computed using the length-volume method (**Equation 3**). This method treats the length of the LV’s axis from base-to-apex as a diameter, and then divides the measured LV volume by the volume of a sphere with that diameter^2,3^.

**Equation 3 (sphericity index, length-volume method)**

$$\frac{LV volume}{Volume of sphere with diameter equal to the LV long-axis length}$$

The Wadell approach from **Equation 2** yields sphericity values that are bounded between 0-1, because a sphere represents the maximum possible volume for a given surface area. The length-volume method from **Equation 3** is bounded only when the base-to-apex dimension is the largest internal dimension in the LV blood pool.

#### Deep learning for semantic segmentation from MRI

##### Model architecture

Multiple segmentation models were trained, but the same model architecture was used for all segmentation models. The model encoder was built from the PyTorch function *mvit_v2_s* (“multiscale vision transformer v2”)^4,5^, and it was paired with a 3D UNet decoder^6^. The output of the deepest layer of the encoder was progressively upsampled in the decoder using 3D convolutional layers, incorporating skip connections from early, intermediate, and deep stages of the decoder; since each imaging view is planar, these models incorporate adjacent temporal data as the third dimension. To match with the pre-trained weights for the encoder described below, the model was built to require video inputs shaped [3, 16, 224, 224], where 3 is the number of color channels, 16 is the number of time points, and the height and width are both 224 pixels.

The encoder stage used weights pre-trained on the DeepMind kinetics dataset that were available in the PyTorch 2.5 *torchvision* package^5,7,8^. The decoder stage used weights that were randomly initialized.

##### Model training

Four deep learning models were trained to perform semantic segmentation (pixel labeling), one per imaging view: the LV short axis (SAX), and the two-, three-, and four-chamber long-axis views (2ch, 3ch, 4ch). All models were built with PyTorch 2.5^8^. Separate models were trained for the short axis view and for the 2-, 3-, and 4-chamber long-axis views: four models in total.

The intrinsic data shape for each cardiac MRI view was [1, 50, height, width], where 1 was the number of color channels and 50 was the number of time points. To comply with the model architecture's input shape requirements, the images were duplicated to create 3 color channels and the height and width were padded to 224. During training, contiguous blocks of 16 frames were selected from a random starting index, allowing for wraparound between the start and end of the CINE (because cardiac motion is cyclic).

Image augmentation was performed with the Kornia library^9^, which was only applied to the training data (not for the validation steps). These transformations included affine transformation (rotation from -180 to 180 degrees, translation by up to 20% in horizontal and vertical directions, and bilinear scaling from 50% to 200%), which was applied to both the input data and the paired segmentation ground truth data. To mimic imaging intensity variation, gamma correction was randomly varied from 0.65 to 1.5 and a plasma brightness transformation was applied, adding noise up to 30% of the maximum brightness level in a fractal pattern across the images. Pixel values were rescaled to the -1 to 1 range before input into the model. Throughout, augmentation was applied in a video-aware sequential manner (i.e., the same transformations were applied to all 16 timepoints).

The FocalDice loss function from Segment Anything^10^ was used with the default hyperparameters. The AdamW^8^ optimizer was used with the default hyperparameters. The batch size was 8. The maximum learning rate was chosen separately for each model using a learning rate finder^11^. At the start of training, the learning rate was warmed up via linear interpolation from 2e-8 to the maximum learning rate over 1,000 steps (minibatches), and the learning rate was cooled to 2e-8 over 1,000 steps at the end of training. In between, the models were trained for 20,000 steps at a constant learning rate. The model weights from the epoch with the best validation loss were saved and used for inference.

The model-specific learning rates and training sample counts were as follows:
- 2ch: 182 training samples, 21 validation samples, max learning rate 1e-3.
- 3ch: 224 training samples, 34 validation samples, max learning rate 8e-4.
- 4ch: 230 training samples, 26 validation samples, max learning rate 3e-4.
- SAX: 571 training samples, 76 validation samples, max learning rate 2e-3.

##### Model inference

At inference time, each 50-timepoint CINE was sampled in 16-frame batches 5 times. For each of these 5 batches, the initial frames were offset by 10 (starting index: 0, 10, 20, 30, 40) to ensure that each frame was covered at least once; for the 30 frames that were sampled twice, an average of the two samples was taken. Height- and width-padding was added to ensure shape-compatibility with the model architecture, and that padding was removed from the output to ensure that the segmentation output retained pixel-correspondence to the input image shape.

##### Model validation

Internal validation for the segmentation output was performed with held-out test samples. Dice scores, a measure of agreement between the manual segmentations and the model predictions for the LV blood pool and myocardium, are reported for the held-out test sets in **Supplementary Table 1**^12,13^. External validation compared to expert-labeled ground-truth data for the SAX view was performed using data from ACDC^14^.

#### Left ventricular surface reconstruction and measurement

##### Shared coordinate system using DICOM metadata

For one individual at one time point, all planar segmentation data (2-, 3-, and 4-chamber long axis data and the short axis stack) were placed into the same 3D coordinate system in *Open3D v0.18.0*, using the DICOM metadata (DICOM tags [0020,0032] and [0020,0037]).

##### Perimeter extraction

Using *scikit-image v0.22.0*, the perimeter of each left ventricular segmentation mask was extracted, resulting in a sparse point cloud representing the LV blood pool surface. To reduce noise, outlier points beyond 3 interquartile ranges from the median distance to the point cloud centroid were removed. This yielded a single, sparse point cloud from each of the short-axis slices and the 2-, 3-, and 4-chamber long-axis slices.

##### Registration and correction of breath-hold errors

Rigid registration to bring the short-axis stack into alignment with the long-axis data was necessary to reduce breath-hold alignment errors. The sampling of points from the long-axis data in the left ventricular blood pool was not uniform. To create a surface with more uniformly spaced points for the registration step, a low-resolution screened Poisson surface reconstruction step^15,16^ was performed using *Open3D* *create_from_point_cloud_poisson* (octree depth 4, scale 2.2), built from the long-axis segmentation data. The vertices of the resulting mesh functioned as a more uniform sparse point cloud, which was used to guide the translation of each short-axis slice only within its own plane (orthogonal to the long-axis) using iterative closest points^17^. This initial long-axis Poisson reconstruction was only used for this registration step; subsequent steps used the originally extracted point clouds.

##### Removal of apical short-axis planes

Because of the tapering of the blood pool near the apex, few pixels from the short-axis slices belong to the LV blood pool at that region and the segmentation at that level was prone to error. Even when accurate, the apical slices contributed only trivially to the volume estimates. Therefore, data from short-axis planes that were within the apical-most 5mm of the LV blood pool as defined by the long axis data were excluded, which then allowed the reconstruction algorithm to use the long-axis data to define the surface boundary near the apex.

##### Removal of short-axis planes basal to the atrioventricular boundary

The *svm.SVC* function from *scikit-learn v1.4.0* was used to estimate a planar boundary between the left atrial segmentation and LV segmentation using the long-axis data. This boundary was used to detect inaccurate LV blood pool segmentation at the base of the heart (namely, due to over-segmentation into the left atrial blood pool). Short axis LV segmentation data on the atrial side of this boundary were discarded prior to volumetric reconstruction.

##### Screened Poisson surface reconstruction and measurement

Using the aligned short- and long-axis sparse surface point clouds as input, screened Poisson surface reconstruction^15,16^ was performed with the *Open3D* *create_from_point_cloud_poisson* function using an octree depth of 5 and a scale parameter of 2.2. The reconstructed mesh was converted to *PyVista* format (*v0.43.2*) and *pymeshfix v0.16.2* was applied, followed by mesh smoothing (with the *PyVista smooth_taubin* function).

The built-in *PyVista* *volume* and *area* functions were called on the output mesh, returning measurements with units of microliters and square millimeters, respectively. These volume and surface area measurements were used to calculate the sphericity measurements as detailed above and the standard LV measurements as detailed in the main **Methods**. The LV long axis length was measured by projecting all long-axis points onto a line normal to the short axis stack.

##### LV mass measurement from surface reconstruction

In addition to the surface reconstruction of the LV blood pool as described above, an additional reconstruction was performed by first merging the segmentation data for the LV blood pool and the LV myocardium. Reconstruction followed the same steps as outlined above for the LV blood pool, producing a combined volume bounded by the outer surface of the LV myocardium. The LV blood pool volume was subtracted from the combined volume, yielding a measurement of myocardial volume at each of the 50 time points during the cardiac cycle for each participant. The myocardial volume was multiplied by 1.05 g/cm^3^ to yield a measurement of LV mass^18^.

#### Technical quality control

3D reconstruction data were analyzed from all individuals who had not withdrawn from the UK Biobank as of the cutoff date described in the **Methods**. Several technical quality control techniques were developed, as described below.

##### Euler characteristic

*PyVista* built-in functions were used to count the number of vertices, edges, and faces of the reconstructed LV blood pool surface mesh. The Euler characteristic was computed as the number of vertices plus the number of edges minus the number of vertices. An Euler characteristic of 2 represents a closed sphere while a number of 0 represents a torus. Individuals who had an Euler characteristic inconsistent with a closed sphere at ventricular end-systole or end-diastole were excluded from all measurements. Because of the sensitivity of sphericity to the shape of the blood pool surface, sphericity was set to a missing value for individuals who had an Euler characteristic below 2 for more than 5% of the time points during the cardiac cycle.

##### High amplitude transient motion

Artifacts causing inaccurate surface reconstructions were noted to occur sporadically, rather than persisting through the cardiac cycle, typically driven by incorrect short-axis segmentation (e.g., segmenting the RV, stomach, or pericardial adipose as left ventricular blood pool). This prompted exclusion of data based on apparent abrupt motion of the centroid of the LV within the short-axis plane orthogonal to the long-axis of the heart: participants were excluded if the location of the centroid of the LV blood pool moved in-plane by more than 4.5mm between any two adjacent timepoints (a duration typically lasting approximately 0.02 second).

##### Gating artifacts

The same multiscale vision transformer (MViTv2) architecture that was used as the encoder for the semantic segmentation models was used to detect visual artifacts typically associated with errors in electrocardiographic gating driven by T-wave abnormalities or arrhythmias in the long-axis images. A training set was generated from manual review by a cardiologist (J.P.P.) where CINEs in which the heart appeared to beat twice, or where the heart appeared to blur, were labeled as ‘artifact’; CINEs with normal motion were labeled as ‘normal’. In total, 1,736 samples were used for training and 220 for validation. After inference, measurements from participants flagged as having gating artifacts in at least two out of the three long-axis planes were excluded.

#### Deep learning model for categorizing atrial motion

During an irregular heart beat, atrial contraction can be absent, which will reduce LV filling and influence parameters such as volume, ejection fraction, and sphericity despite being extrinsic to the LV itself. No electrocardiographic signal data were available from the moment of the MRI to directly identify arrhythmia; therefore, assessment of atrial motion was made based on imaging. Atrial motion was categorized as either standard triphasic (reservoir, conduit, and atrial kick) or biphasic (absent atrial kick). To make these assignments, a deep learning model was developed.

To generate a training set enriched for biphasic motion, the UK Biobank 12-lead ECG data, which was obtained during the same assessment center visit as the imaging data, were used to identify individuals with and without atrial fibrillation. (Because arrhythmia is not permanent for most individuals, the presence of atrial fibrillation on ECG can enrich for biphasic motion during imaging, but does not guarantee its presence.) Visual categorization of cardiac motion was made by a cardiologist (J.P.P.) for 1,915 CINEs: 815 from 2ch, 544 from 3ch, and 556 from 4ch. Model training was performed using data from 1,557 training samples and 207 validation samples (with training/validation splits based on the participant, to avoid any individual contributing data both to the training and the validation sets).

The model architecture was fine-tuned from the same pre-trained MViTv2 architecture that was used as the encoder for the segmentation models described above^4,5^, adapted with a linear layer to emit predictions for biphasic and triphasic motion. For this model, the data representation was condensed: each of 3 adjacent timepoints were treated as color channels, and timepoints 24 and 25 were dropped, allowing the remaining 48 frames to be represented in one 16-timepoint 3-channel object. During model training, the AdamW^8^ optimizer was used with the default hyperparameters. The loss function was cross entropy. The batch size was 32. The maximum learning rate (here, 2e-5) was chosen using a learning rate finder^11^. At the start of training, the learning rate was warmed up via linear interpolation from 2e-8 to 2e-5 over 1,000 steps, and the learning rate was cooled to 2e-8 over 1,000 steps at the end of training. In between, the model was trained for 10,000 steps at the maximum learning rate (2e-5). The model weights from the epoch with the best validation loss were saved and used for inference in all participants.

After inference, performance was assessed in data from 151 held-out test samples: of the 39 triphasic and 112 biphasic CINEs in the test set, the model had an accuracy of 0.993, incorrectly labeling one biphasic CINE as triphasic (sensitivity 0.991, specificity 1.000, positive predictive value 1.000, negative predictive value 0.975).

#### Wasserstein distance

While it is common to test for normality with the Kolmogorov-Smirnov test or the Anderson-Darling test, even minor deviations from the normal distribution will be statistically significant with a sample size in the tens of thousands. Therefore, rather than performing null hypothesis testing, the Wasserstein-1 (“earth mover’s”) distance was used to give a sense of distributional similarity. The Wasserstein distance represents how much one distribution would need to shift to match another distribution, in the same units as the trait itself. This approach has been used for analyzing anthropometric traits previously^19^.

#### UK Biobank whole genome sequencing

All genetic analyses were conducted with respect to GRCh38 using whole genome sequencing (WGS) data, which were available for 490,036 UK Biobank participants after central quality control^20^. Germline variant calling had been performed using the DRAGEN v3.7.8 germline pipeline^21^. Then, the DRAGEN MLR v4.2.4 software was run on the sample level to recalibrate the variant quality. A recalibrated joint call set was produced with the DRAGEN IGG pipeline v4.2.4, and variants with recalibrated QUAL < 3 were removed prior to aggregation, yielding 1.1 billion variant sites (1.5 billion alleles). Multiallelic sites were centrally split into pseudobiallelic sites. Pseudobiallelic variants with QUAL < 0.1 excluded. Variants were assembled into Plink2-formatted files^22^, available from UK Biobank field ID #24308. All variants with a non-PASS filter were excluded before analysis in the present study, and all pseudobiallelic variant names were replaced with “DRAGEN:chr:pos:ref:alt” to facilitate identifiability.

#### Genetic analysis sample quality control

For genetic analyses, participants were excluded due to a history of cardiac transplantation, aortic valve procedures, other cardiac surgery, anterior MI, or ischemic cardiomyopathy. Participants were also excluded due to having no genetic data that passed whole-genome-sequencing quality control, sex chromosome aneuploidy, or being an outlier for heterozygosity or relatedness.

#### Approximate power boundary for common- and low-frequency variants

For visualization, approximate statistical power boundaries were computed for autosomes and chromosome X, showing the expected detectable effect size (in trait-SD units). For each minor-allele frequency, the minimum additive effect that would attain two-sided genome-wide significance (at alpha = 5E-08) with 50% power in a cohort of 80,000 individuals was solved for. Power was computed using the quantitative-trait noncentrality parameter from Sham and Purcell^23^. These values differ for autosomes and the X chromosome because men are hemizygous for X and coded as 0/2 in REGENIE, permitting the detection of smaller effect sizes at the same power threshold in a sex-balanced cohort.

#### HapMap3+ LD matrix generation for PRScs

Linkage disequilibrium (LD) matrices were generated for the 1,408,687-million variant panel described in the *ldpred2* manuscript as “HapMap3+”^24^. PRScs requires dense LD matrices within each block; however, the *ldpred2* author-provided HapMap3+ LD matrices are sparse. Therefore, LD was recomputed for these variants using the UK Biobank WGS data with plink2^22^ within GRCh38-specific LD blocks, derived from the EUR 1000-Genomes participants with *pyrho*^25,26^.

#### Polygenic score residualization using 1000 Genomes

To reduce ancestry bias in the point estimates of the polygenic scores, before testing for downstream association, polygenic scores were residualized using data from the 1000 Genomes Project. The 1000 Genomes Phase III WGS data aligned to GRCh38 produced by Byrska-Bishop and made available in .pgen format by the plink2 authors were used for this step ^22,27^. The polygenic scores for each LV measurement were applied, and then the first 20 principal components (PCs) of ancestry were used to estimate the score in a linear model (step 1). The squared residual values were then predicted from the first 20 PCs (step 2). Finally, each residual from step 1 was divided by the square-root of the variance predicted in step 2 to obtain a mean- and variance-corrected polygenic score.

These formulas were then applied to the other cohorts (UK Biobank and *All of Us*), yielding mean- and variance-corrected polygenic scores, following the approach of Khan, *et al*^28^. Consequently, the polygenic score values have an absolute interpretation as units of 1 standard deviation within the 1000 Genomes Project, although the scores were scaled to mean 0 and standard deviation 1 (or cut at quantile thresholds) within the respective analytic cohorts for all analyses.

#### Initial approach with single-frame segmentation

An initial draft of this work was undertaken using single-image segmentation models, described in this section. These draft models were built with PyTorch^8^. All draft models used the pre-built ConvNext library available in PyTorch 2.0’s *torchvision* package^8,29^. The 4ch draft model was trained first and used the ConvNext-Small architecture, while all subsequent draft models (2ch, 3ch, and SAX) were trained using the larger ConvNext-Base architecture. To create a U-Net for semantic segmentation (pixel labeling), these draft models were modified to add a symmetric decoding pathway with skip-connections from each downsampling block. All layers were trainable (“unfrozen”) during training.

During training of these draft models, data augmentation was performed with the Kornia library^9^, which was only applied to the training data (not the validation data). These transformations included affine transformation (rotation from -180 to 180 degrees, translation by up to 5%, rescaling from 90% to 110%), which was applied to both the input data and the paired segmentation ground truth data. Gamma was randomly varied from 0.65 to 1.5, and random patches were erased (comprising 2-10% of the image).

For all of these draft models, the loss function was the FocalDice loss from Segment Anything^10^ with the default hyperparameters. The optimizer was AdamW^8^ with a weight decay of 1e-03. The OneCycleLR PyTorch scheduler was used for superconvergence^30,31^. The models were trained for 500 epochs, and the model weights from the epoch with the best validation loss were retained.

The learning rate for each was chosen using a learning rate finder^11^. The batch sizes were generally chosen to fit into the video random access memory (VRAM) of a 16GB VRAM device. The learning rates and other model-specific hyperparameters for the 2ch, 3ch, 4ch, and SAX draft models were as follows:
- 2ch draft: ConvNextBase, 146 training samples, 17 validation samples, batch size 20, min learning rate 1e-5, max learning rate 5e-4.
- 3ch draft: ConvNextBase, 231 training samples, 26 validation samples, batch size 20, min learning rate 1e-5, max learning rate 5e-4.
- 4ch draft: ConvNextSmall, 160 training samples, 38 validation samples, batch size 40, min learning rate 1e-5, max learning rate 2e-3.
- SAX draft: ConvNextBase, 606 training samples, 68 validation samples, batch size 16, min learning rate 1-e5, max learning rate 5e-4.

REGENIE GWAS summary statistics for the standard LV measurements from the first 63,196 UK Biobank MRI participants will be available upon publication at doi:10.5281/zenodo.14025158.

### Supplementary Tables

#### Supplementary Table 1: Semantic segmentation accuracy

| **Validation Set** | **View** | **Structure** | **Samples** | **Dice** | **SD** |
| --- | --- | --- | --- | --- | --- |
| UK Biobank | 2ch | LV blood pool | 21 | 0.94 | 0.04 |
| UK Biobank | 2ch | LV myocardium | 21 | 0.83 | 0.06 |
| UK Biobank | 3ch | LV blood pool | 39 | 0.96 | 0.04 |
| UK Biobank | 3ch | LV myocardium | 39 | 0.85 | 0.06 |
| UK Biobank | 4ch | LV blood pool | 32 | 0.95 | 0.03 |
| UK Biobank | 4ch | LV myocardium | 32 | 0.87 | 0.04 |
| UK Biobank | SAX | LV blood pool | 32 | 0.95 | 0.03 |
| UK Biobank | SAX | LV myocardium | 32 | 0.88 | 0.11 |
| ACDC | SAX | LV blood pool | 132 | 0.92 | 0.08 |
| ACDC | SAX | LV myocardium | 132 | 0.83 | 0.09 |

LV: left ventricular. SD: standard deviation. ACDC: Automated Cardiac Diagnosis Challenge^14^. The Dice score weights each image’s score by its number of pixels matching the respective structure in either the truth labels or the predicted labels.

#### Supplementary Table 2: Approaches for estimating LV sphericity

| **Work** | **Sample size** | **Formula** |
| --- | --- | --- |
| Ambale-Venkatesh, 2017 | MESA; 4,884 | $\frac{LV volume}{{(Length of LV base to apex)}^{3}\times\frac{\pi}{6}}$ |
| Vukadinovic, 2023 | UK Biobank; 38,897 | $\frac{Width of 4ch 2D LV bounding box}{Height of 4ch 2D LV bounding box}$ |
| This work | UK Biobank; 82,674 | $\frac{Surface area of sphere with same volume as LV}{Surface area of 3D LV blood pool}$ |

#### Supplementary Tables 3-5 are in the spreadsheet

#### Supplementary Table 6: Heritability

| Trait | H2G | SE |
| --- | --- | --- |
| LV Sphericity, ED | 0.364 | 0.008 |
| LV Sphericity, ES | 0.348 | 0.008 |
| LVEDV | 0.397 | 0.008 |
| LVESV | 0.396 | 0.008 |
| SV | 0.304 | 0.008 |
| LVEF | 0.314 | 0.008 |
| LV Mass | 0.364 | 0.008 |
| LV Surface Area, ED | 0.405 | 0.008 |
| LV Surface Area, ES | 0.398 | 0.008 |

H2G: Narrow-sense heritability estimated by BOLT-LMM. SE: Standard error. ED: end-diastole. ES: end-systole.

#### Supplementary Tables 7-18 are in the spreadsheet

### Supplementary Figures

#### Supplementary Figure 1: Sample flow diagram

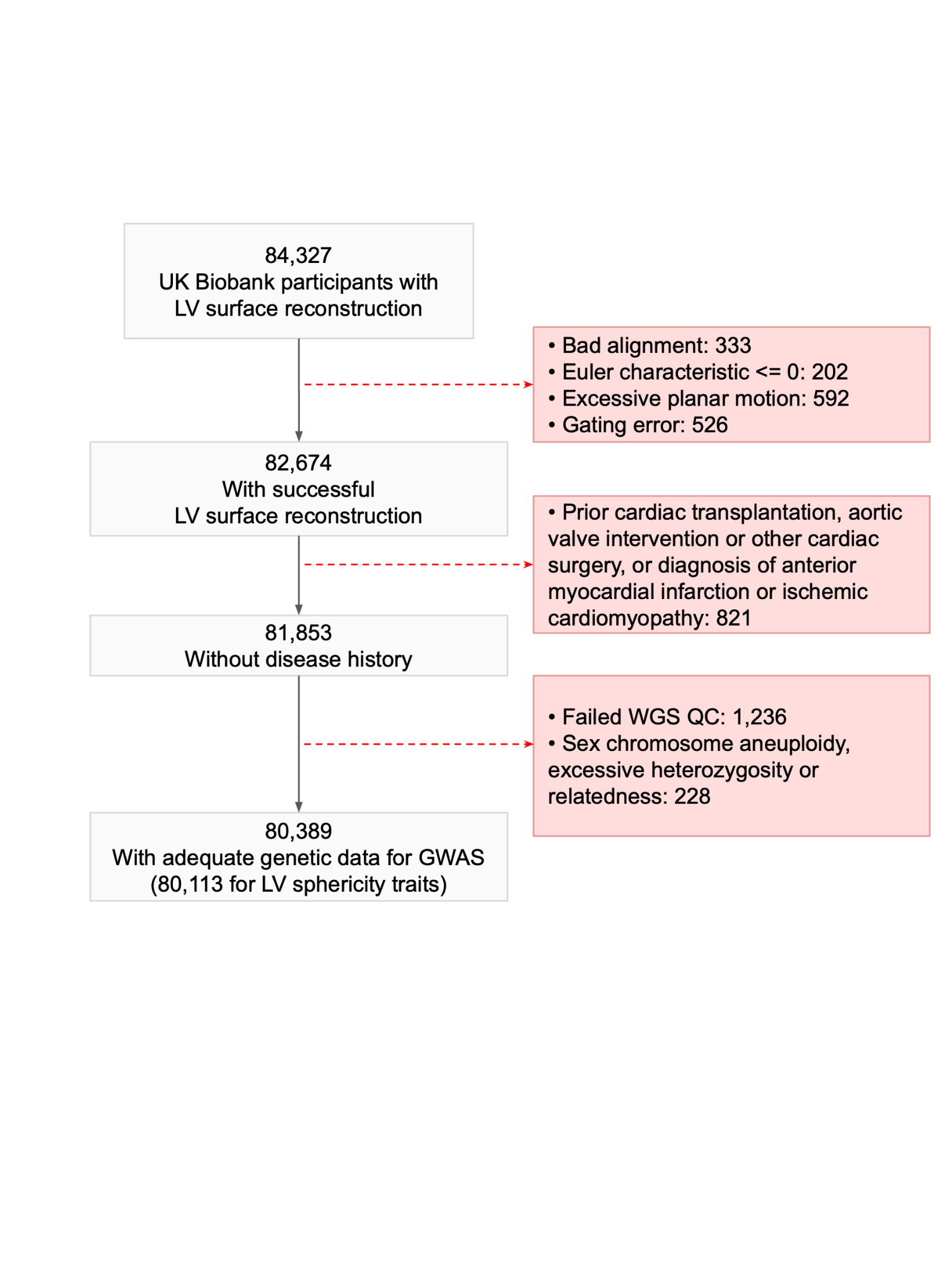

LV: left ventricle. GWAS: genome-wide association study.

#### Supplementary Figure 2: Sex stratified population distribution

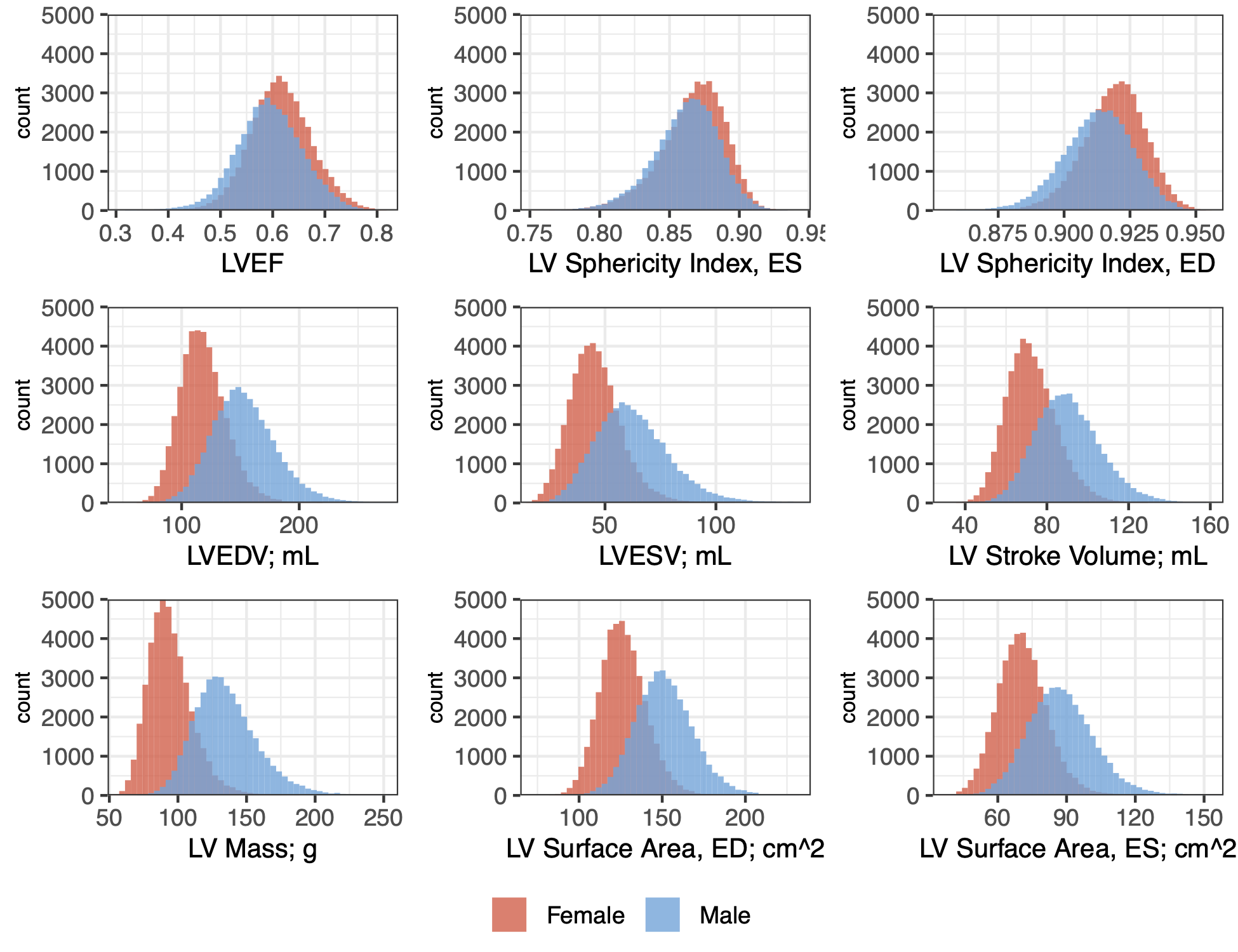

ED: End-diastole. ES: End-systole. LV: left ventricular. LVEDV: LV end-diastolic volume. LVESV: LV end-systolic volume. LVEF: LV ejection fraction. Values beyond 5 standard deviations of the mean are excluded to emphasize the central tendency for display.

#### Supplementary Figure 3: Population distribution of LV sphericity

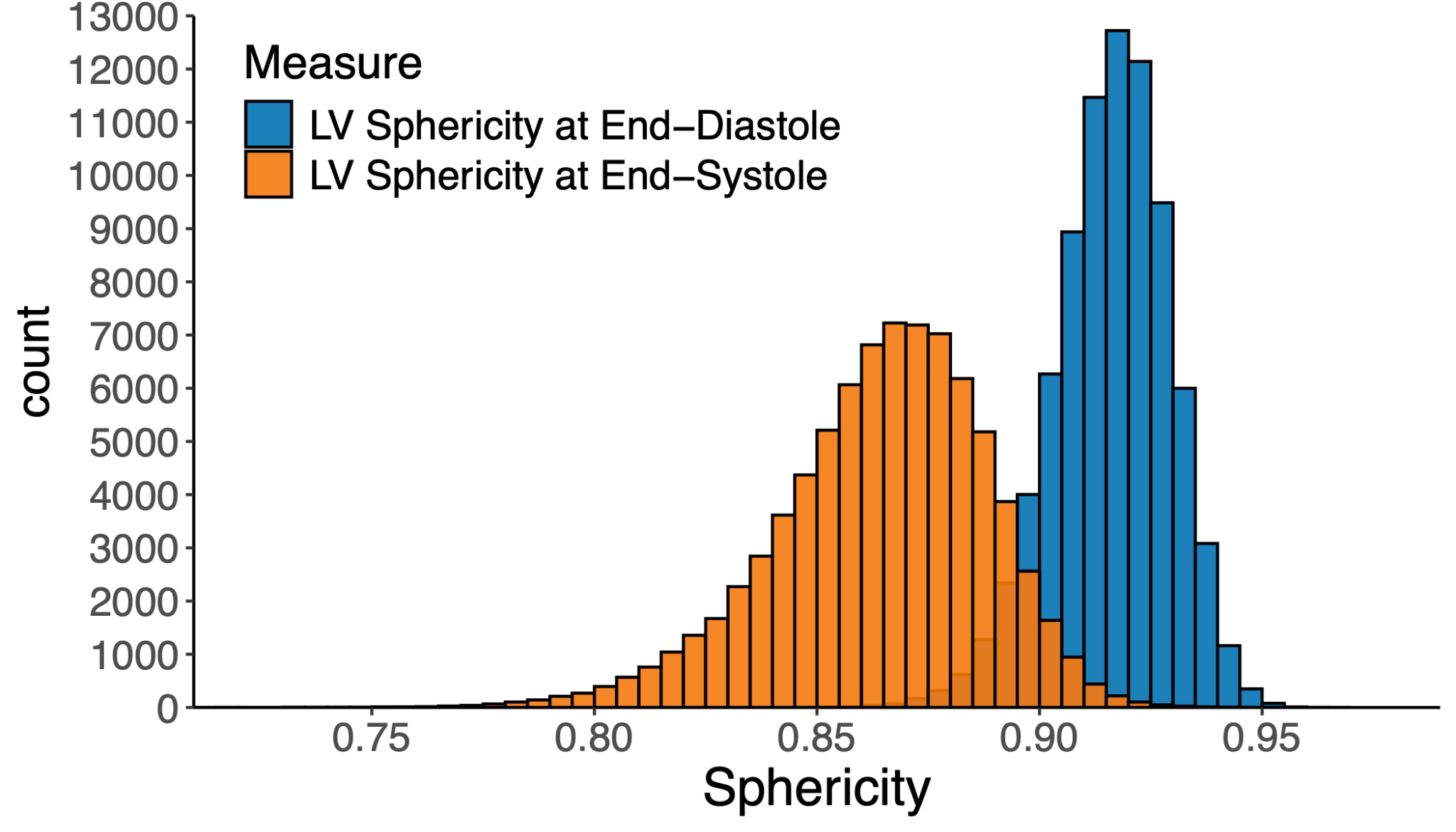

Overlapping regions are shown with partial transparency. LV: left ventricular.

#### Supplementary Figure 4: Correlation between LV traits

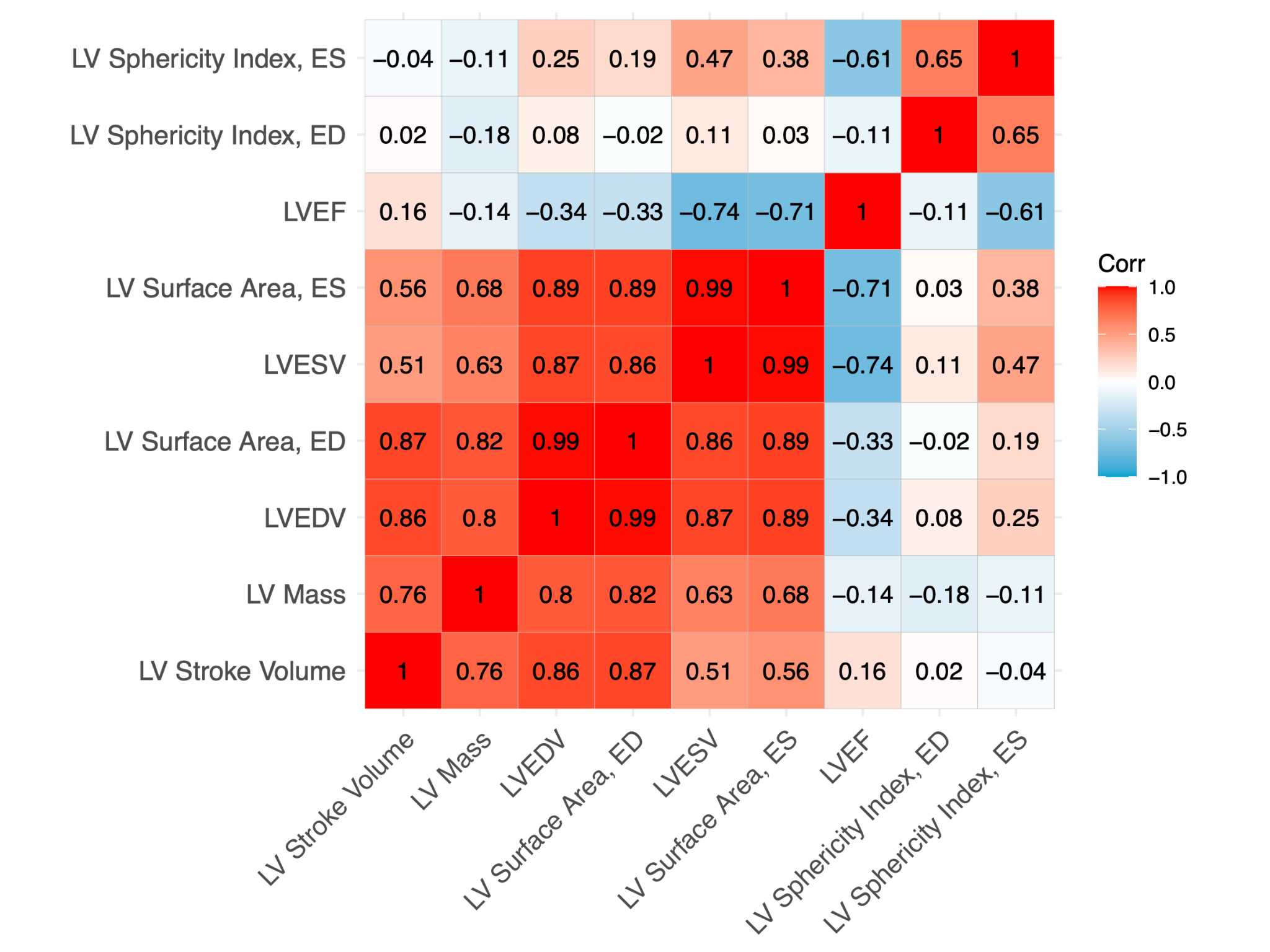

LV: left ventricular. LVEDV: LV end-diastolic volume. LVESV: LV end-systolic volume. LVEF: LV ejection fraction. ED: end-diastole. ES: end-systole.

#### Supplementary Figure 5: Correlation with anthropometric traits

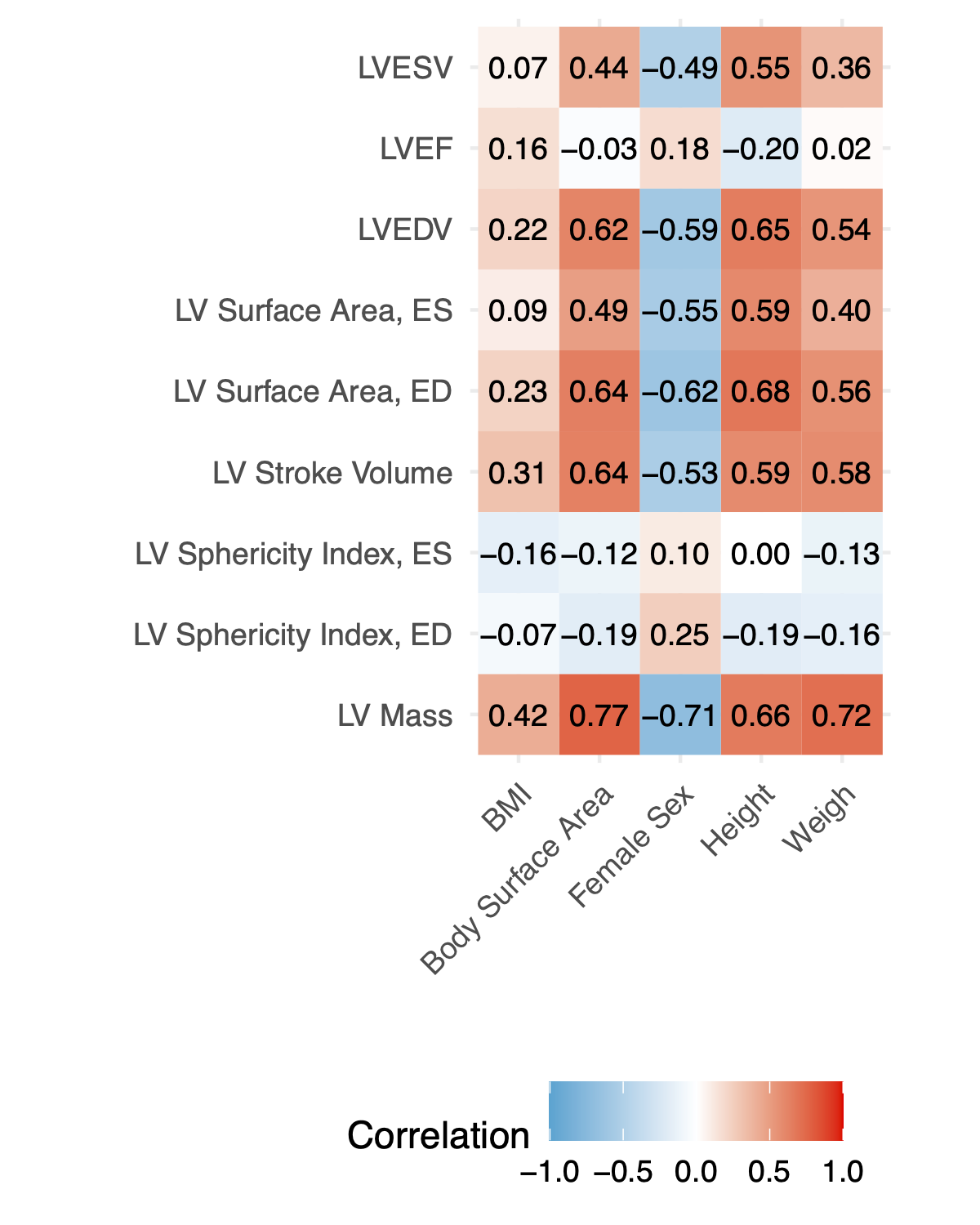

BMI: Body mass index. LV: left ventricular. LVEDV: LV end-diastolic volume. LVESV: LV end-systolic volume. LVEF: LV ejection fraction. ED: end-diastole. ES: end-systole.

#### Supplementary Figure 6: Joint models (LVEF and sphericity for DCM; LVM and sphericity for HCM)

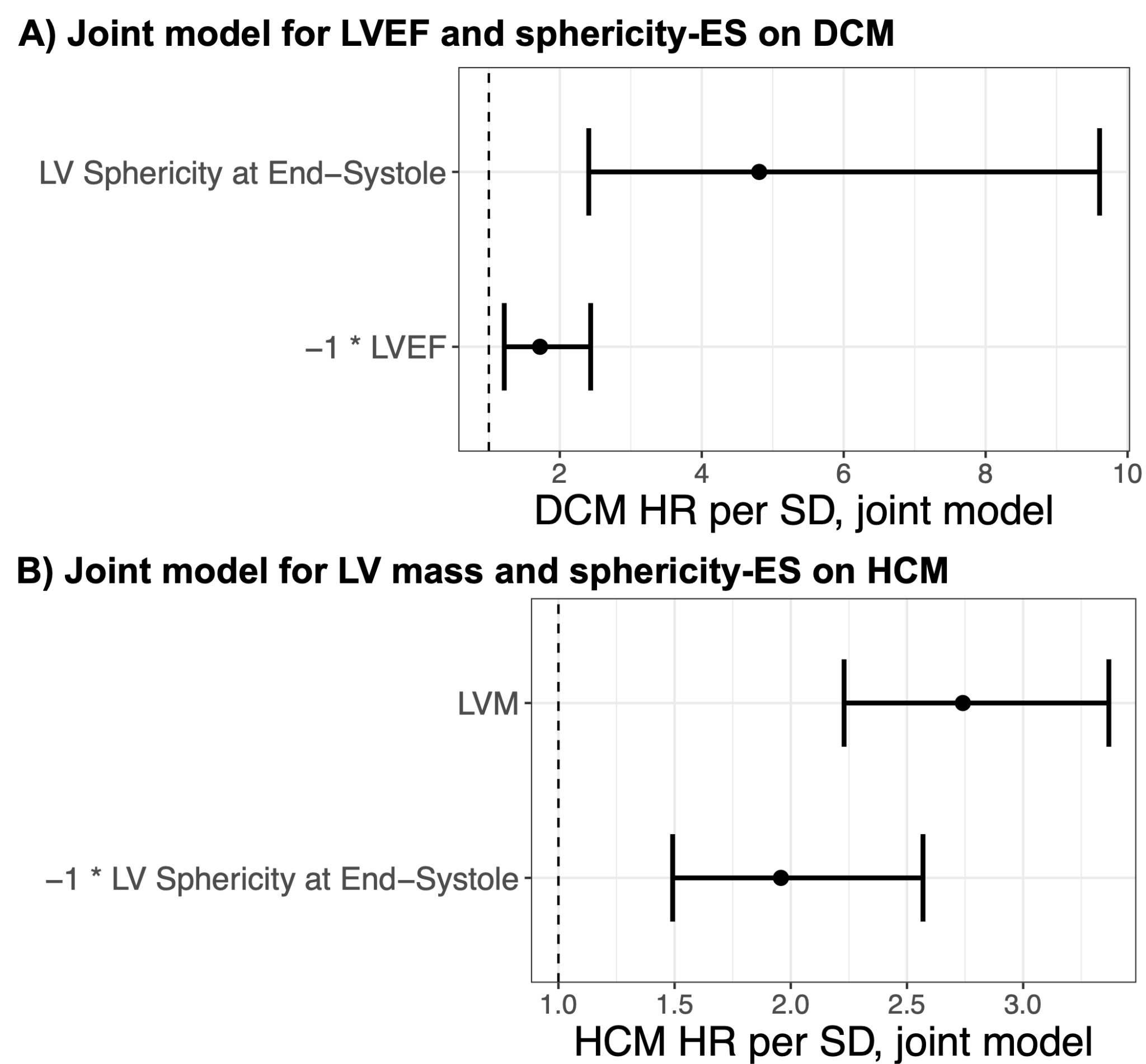

Both models are also adjusted for sex and age at the time of imaging. **Panel A**: Effect estimates for sphericity-ES and LVEF in a Cox model for DCM. The effect estimate for LVEF is multiplied by -1 before exponentiation so that the direction of effect is aligned with the risk-increasing direction to allow for comparison. **Panel B**: Effect estimates for sphericity-ES and LV mass in a Cox model for HCM. The effect estimate for sphericity-ES is multiplied by -1 before exponentiation so that the direction of effect is aligned with the risk-increasing direction to allow for comparison.

#### Supplementary Figure 7: Genetic correlation between LV traits

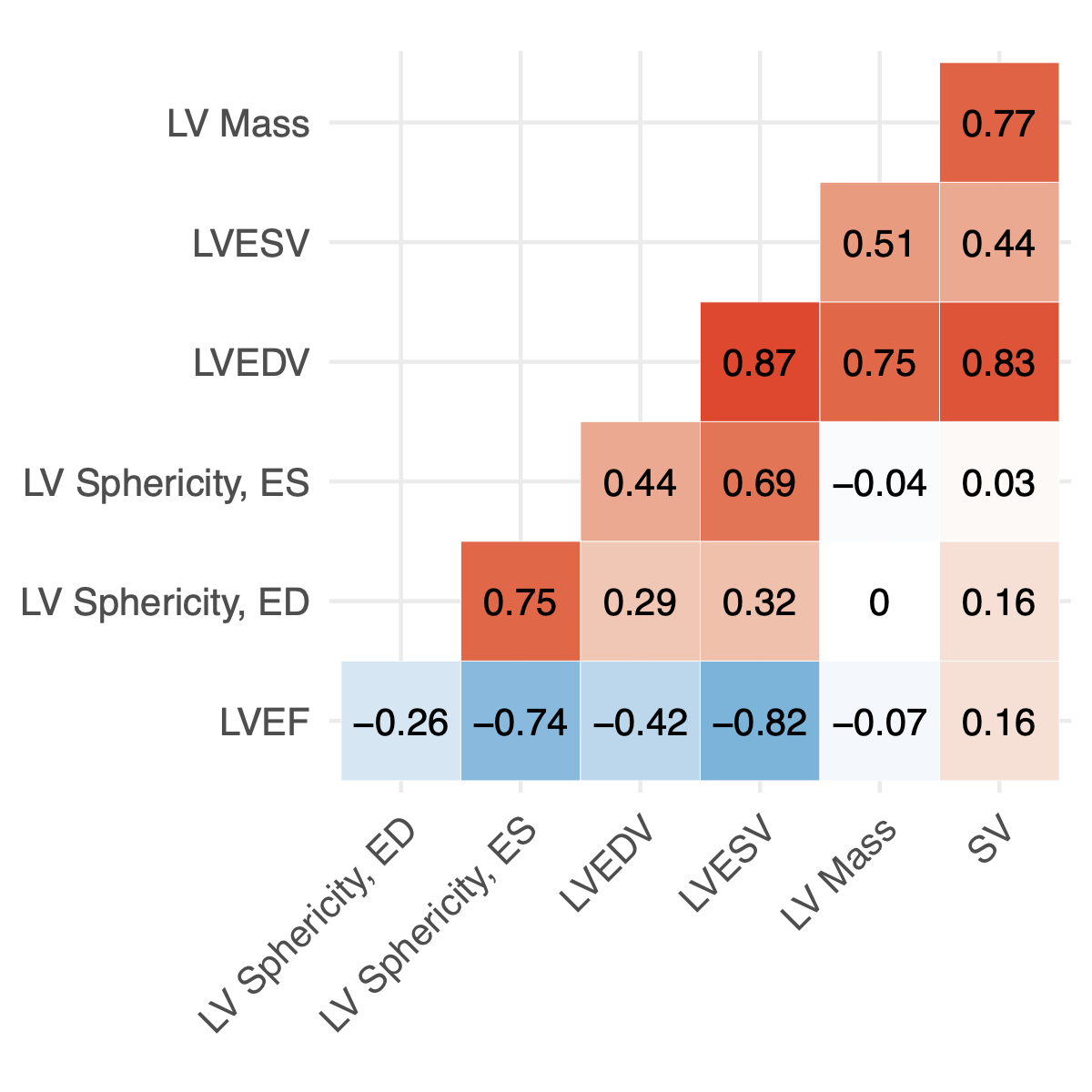

BOLT-REML genetic correlation estimates from UK Biobank GWAS participant data. LV: Left ventricular. LVESV: LV end-systolic volume. LVEDV: LV end-diastolic volume. ES: end systole. ED: end diastole. LVEF: LV ejection fraction. Backing data are available in **STAAA**.

#### Supplementary Figure 8: Manhattan plots for non-sphericity LV traits

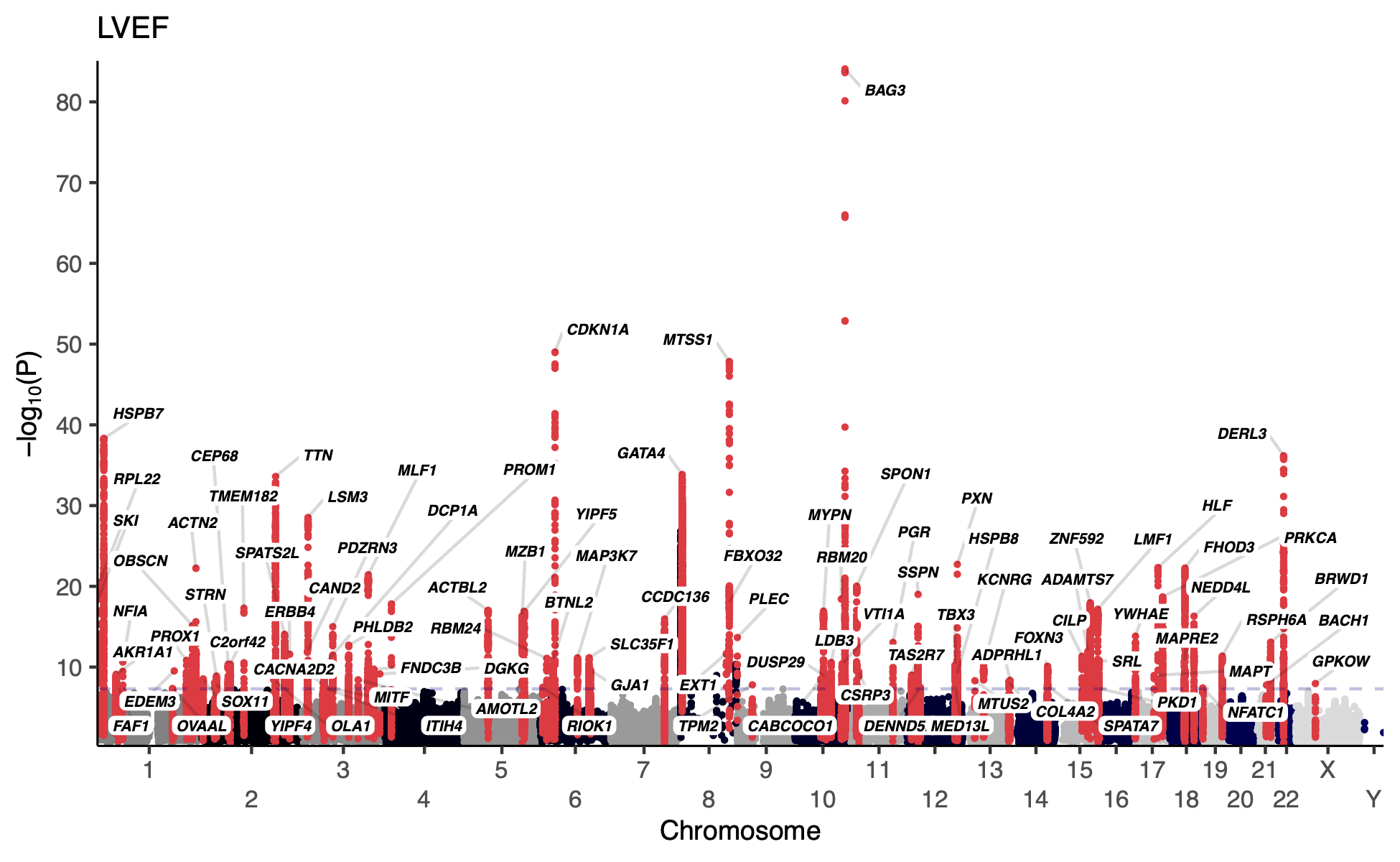

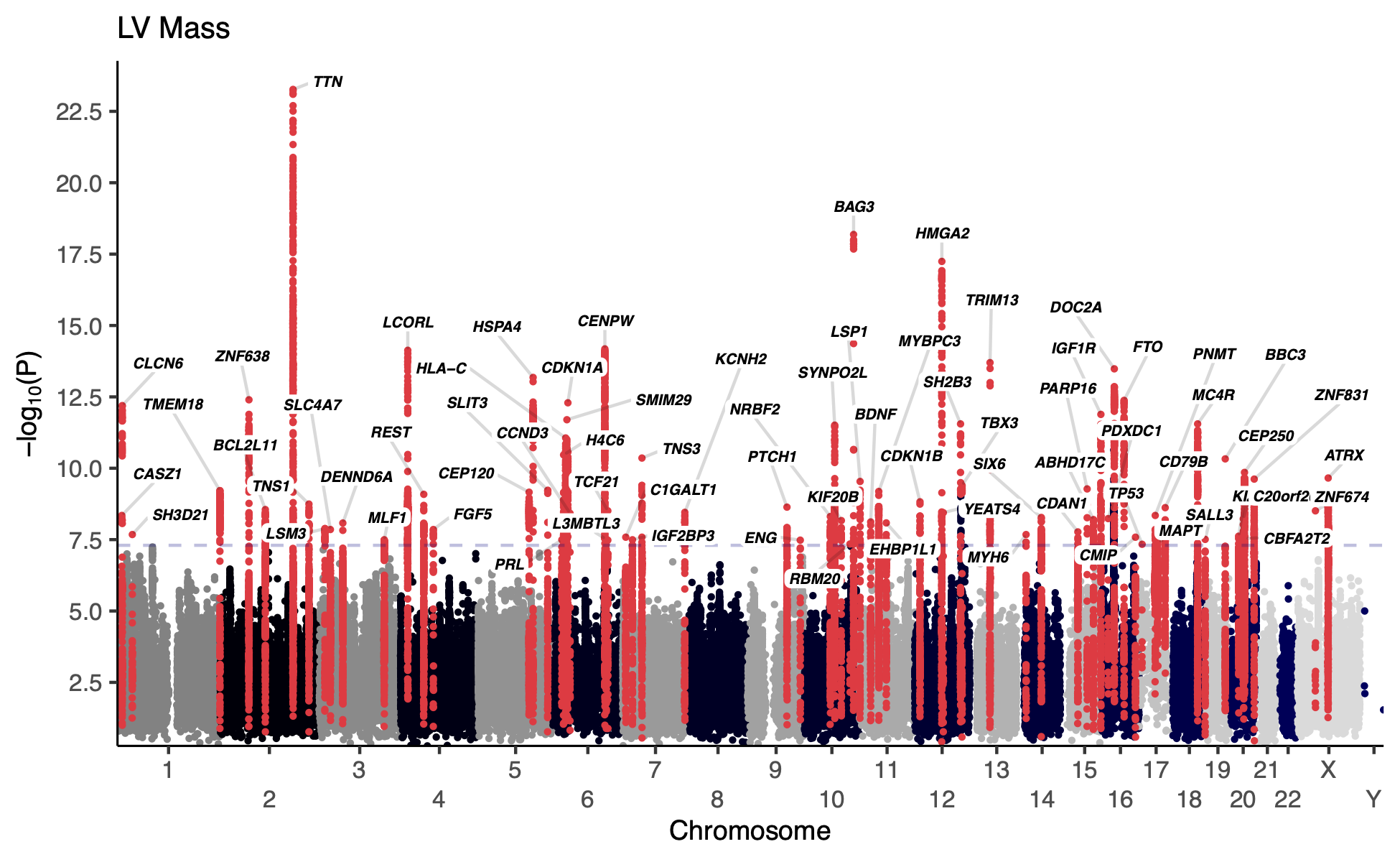

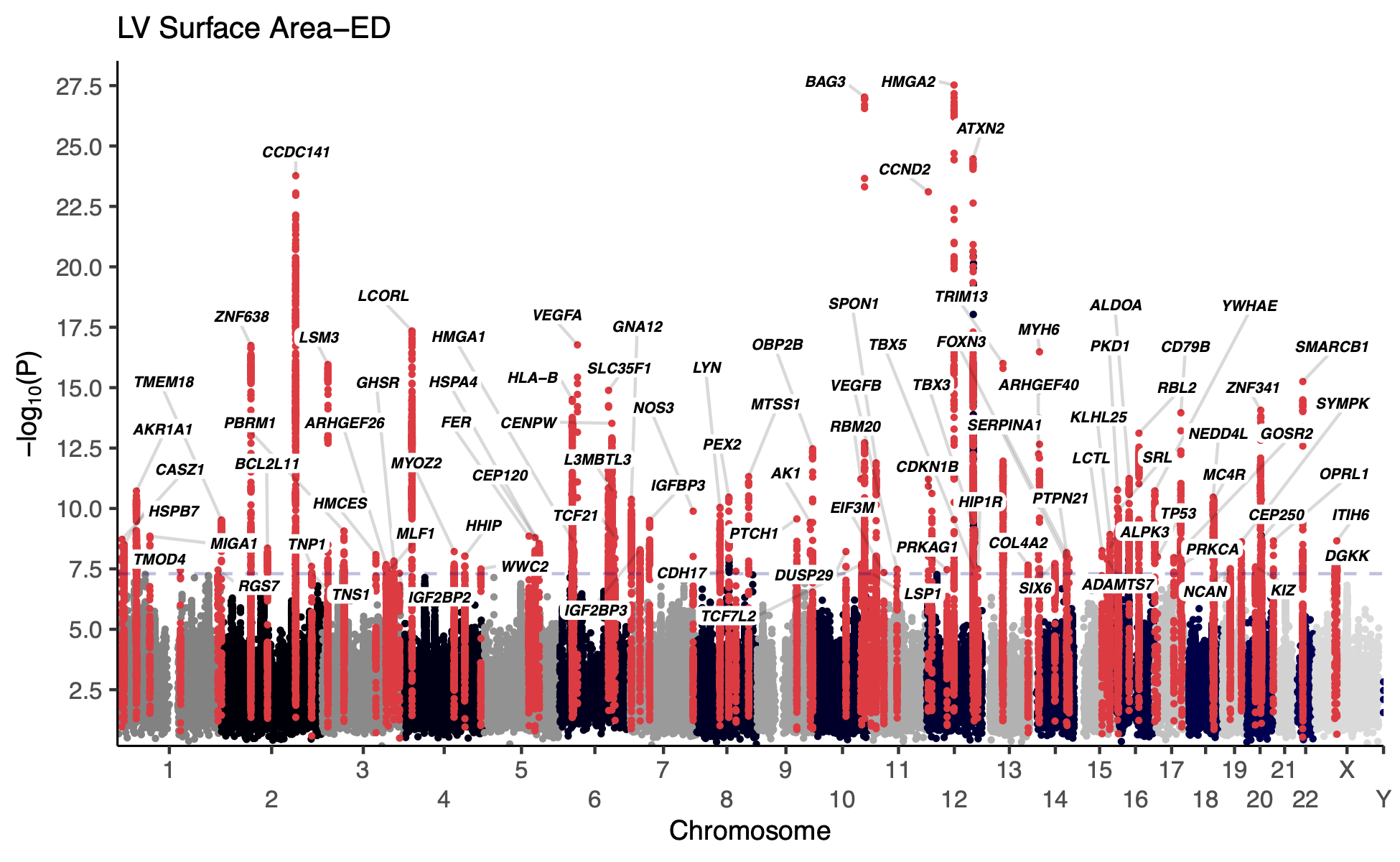

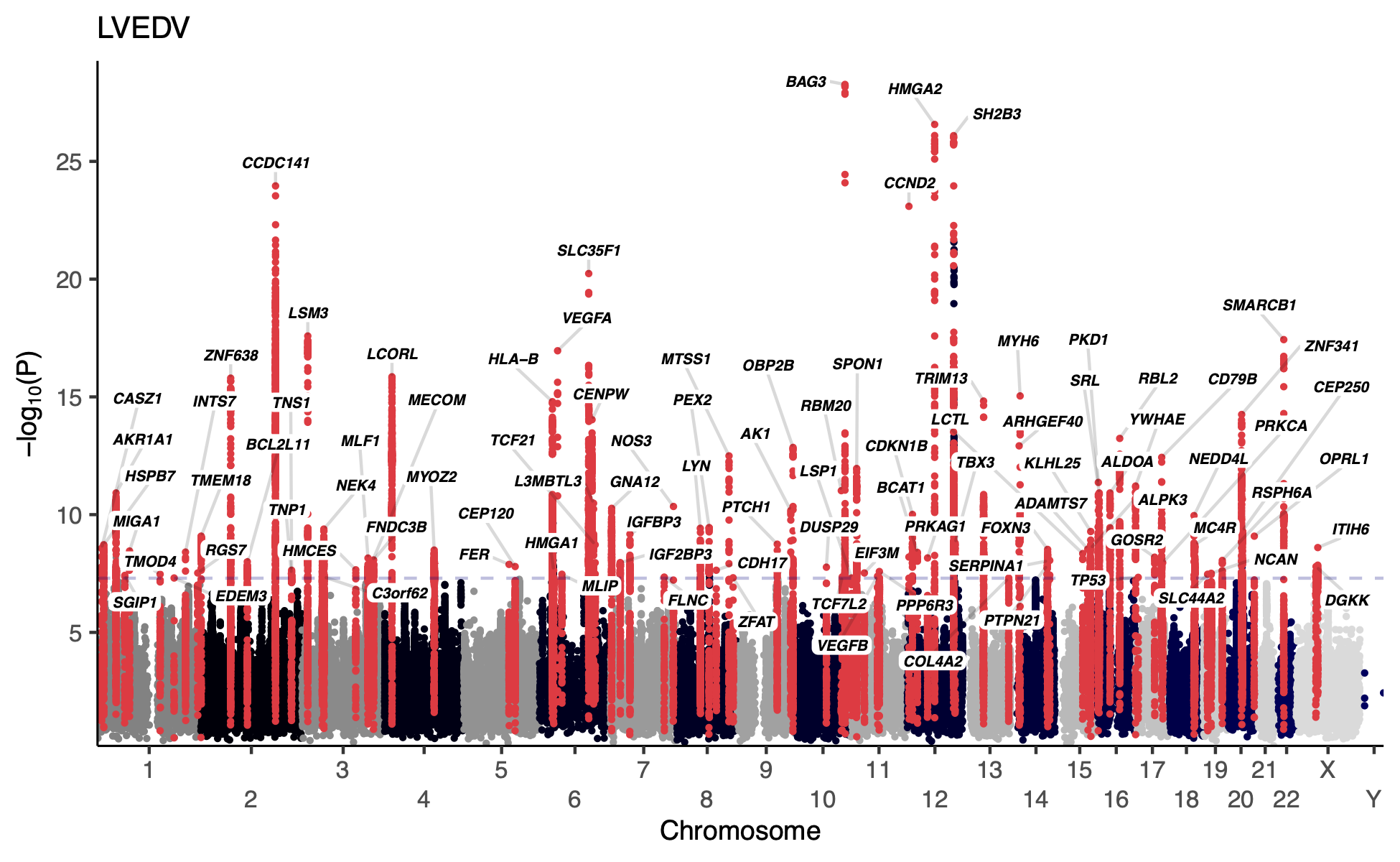

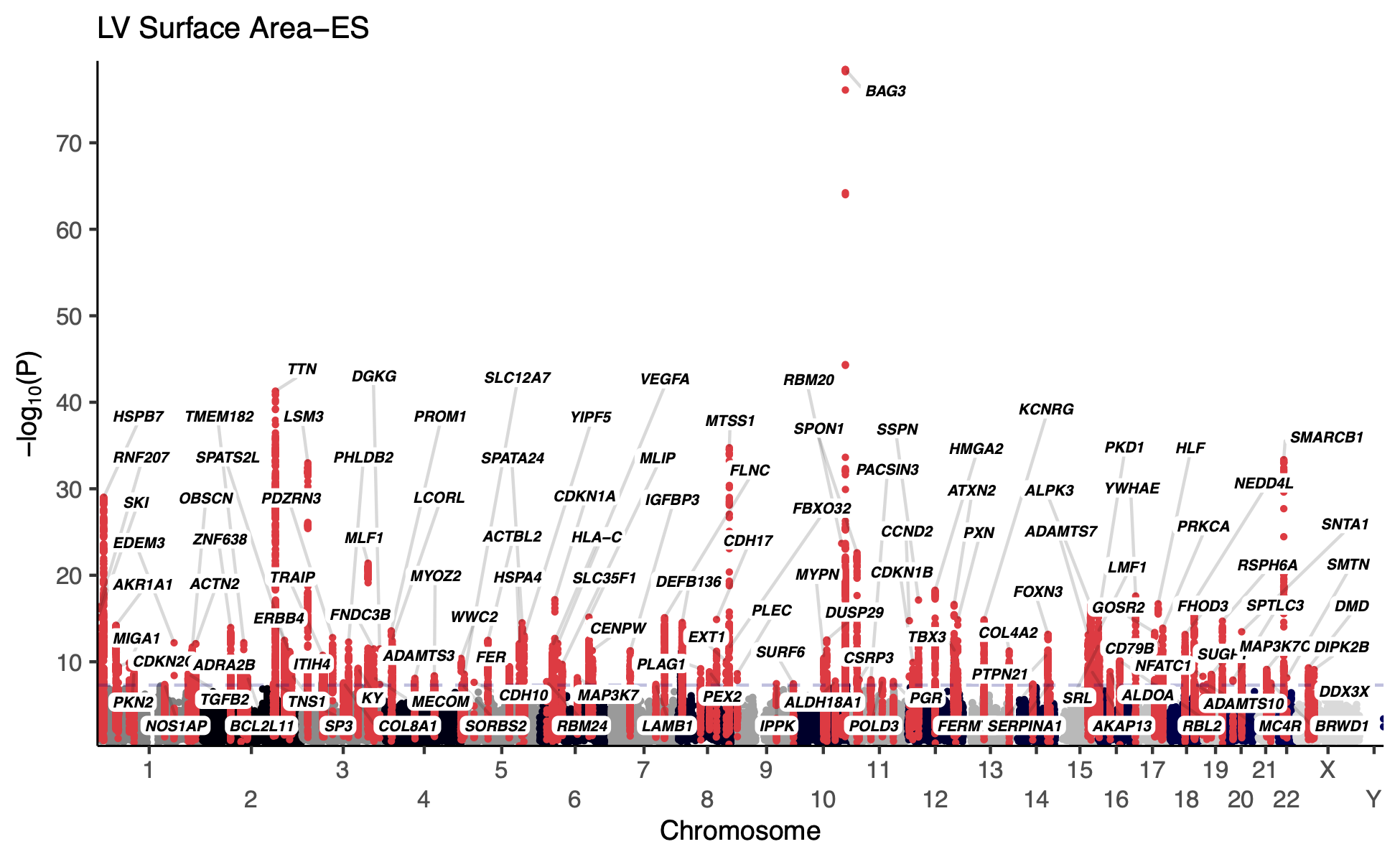

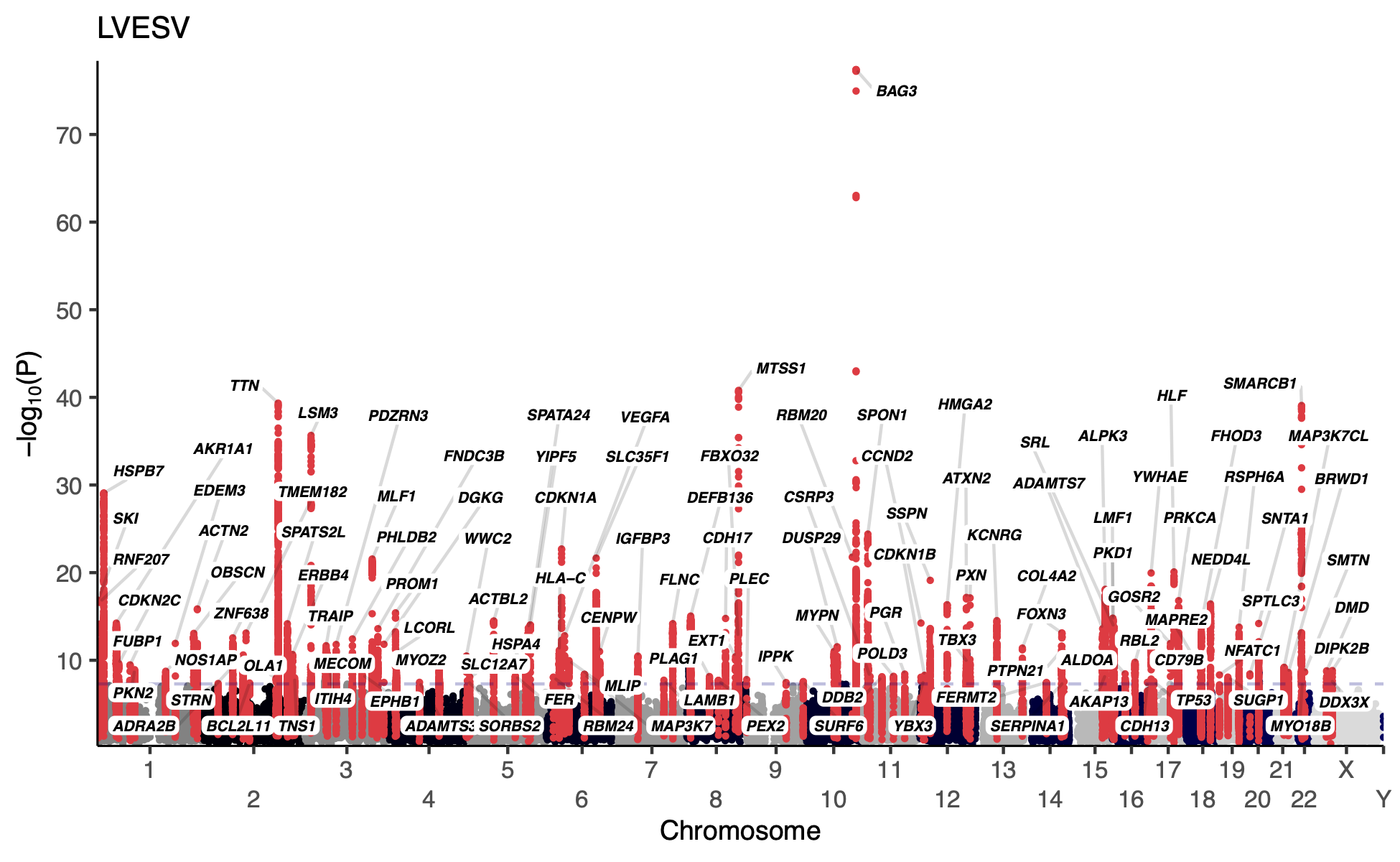

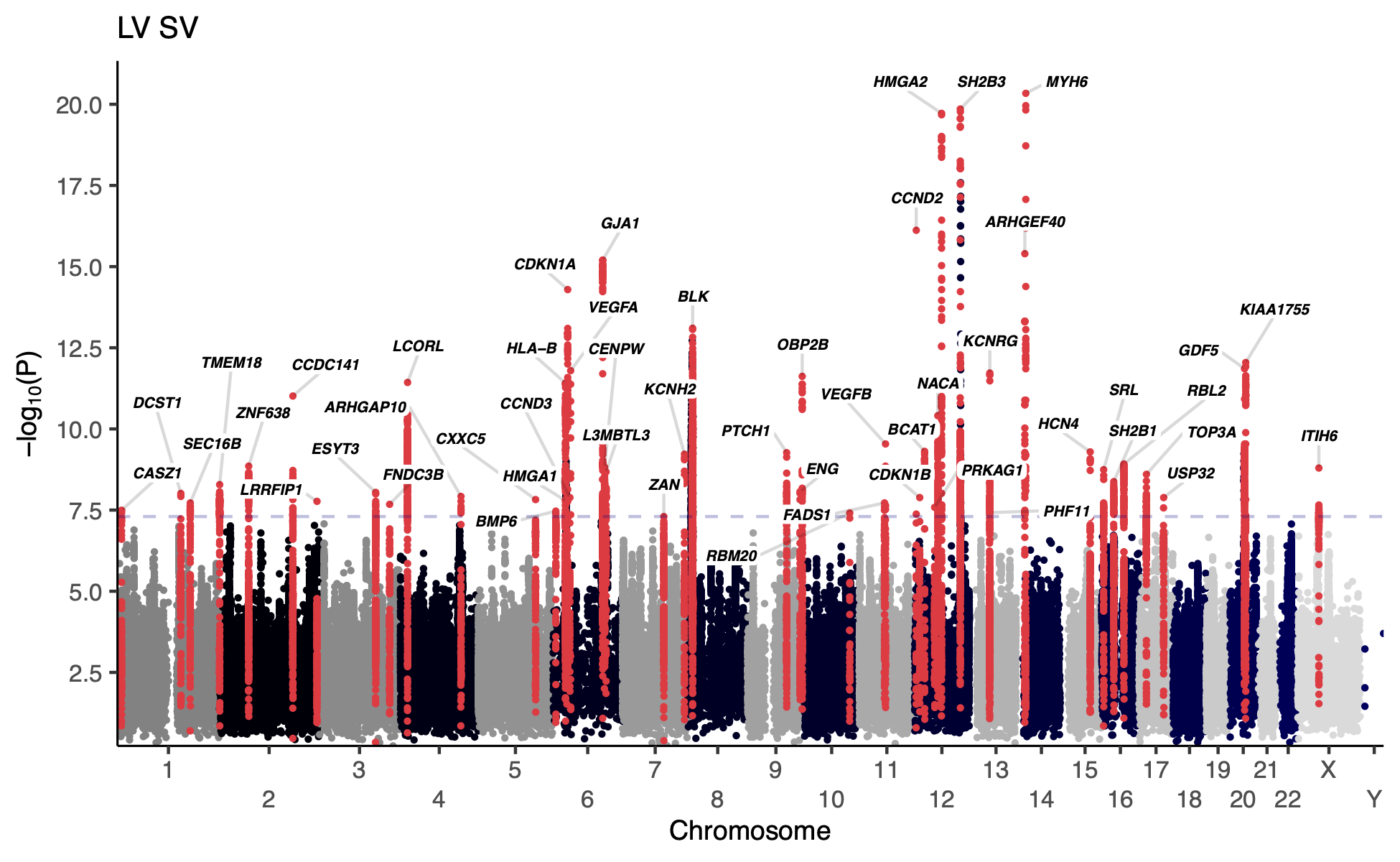

Manhattan plots for non-sphericity LV traits are depicted. Variants at significant loci are depicted in red, with a label identifying the nearest gene to the lead variant at the locus.

#### Supplementary Figure 9: Loci associated only with sphericity

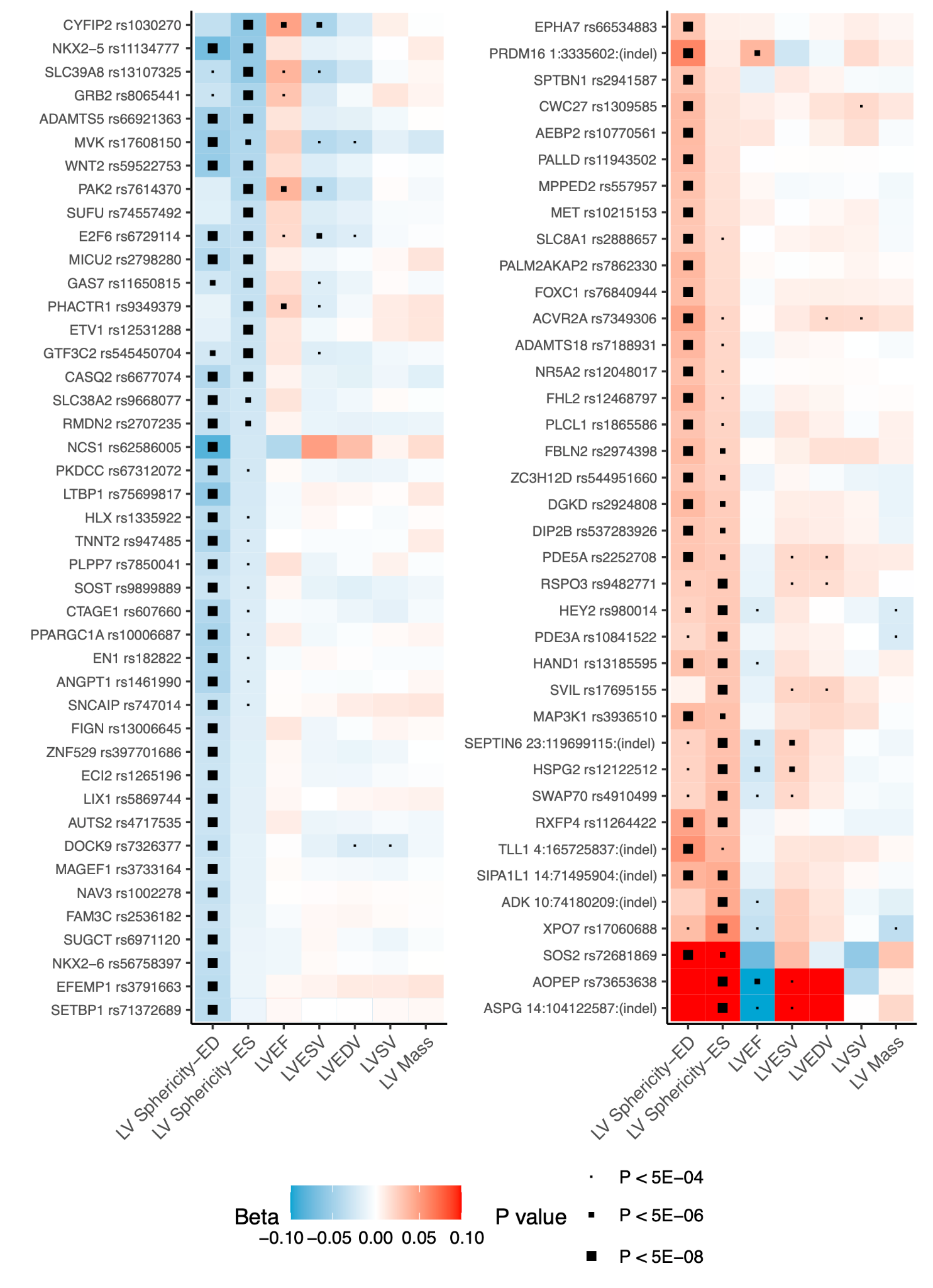

The lead variant and its nearest gene are displayed for all loci that were associated at P<5E-08 for sphericity-ED or sphericity-ES, excluding any variant that was also associated at P<5E-08 for the standard LV measurements. LV: left-ventricular. LVEF: LV ejection fraction. LVESV: LV end-systolic volume. LVEDV: LV end-diastolic volume. LVSV: LV stroke volume.

#### Supplementary Figure 10: GTEx tissue types

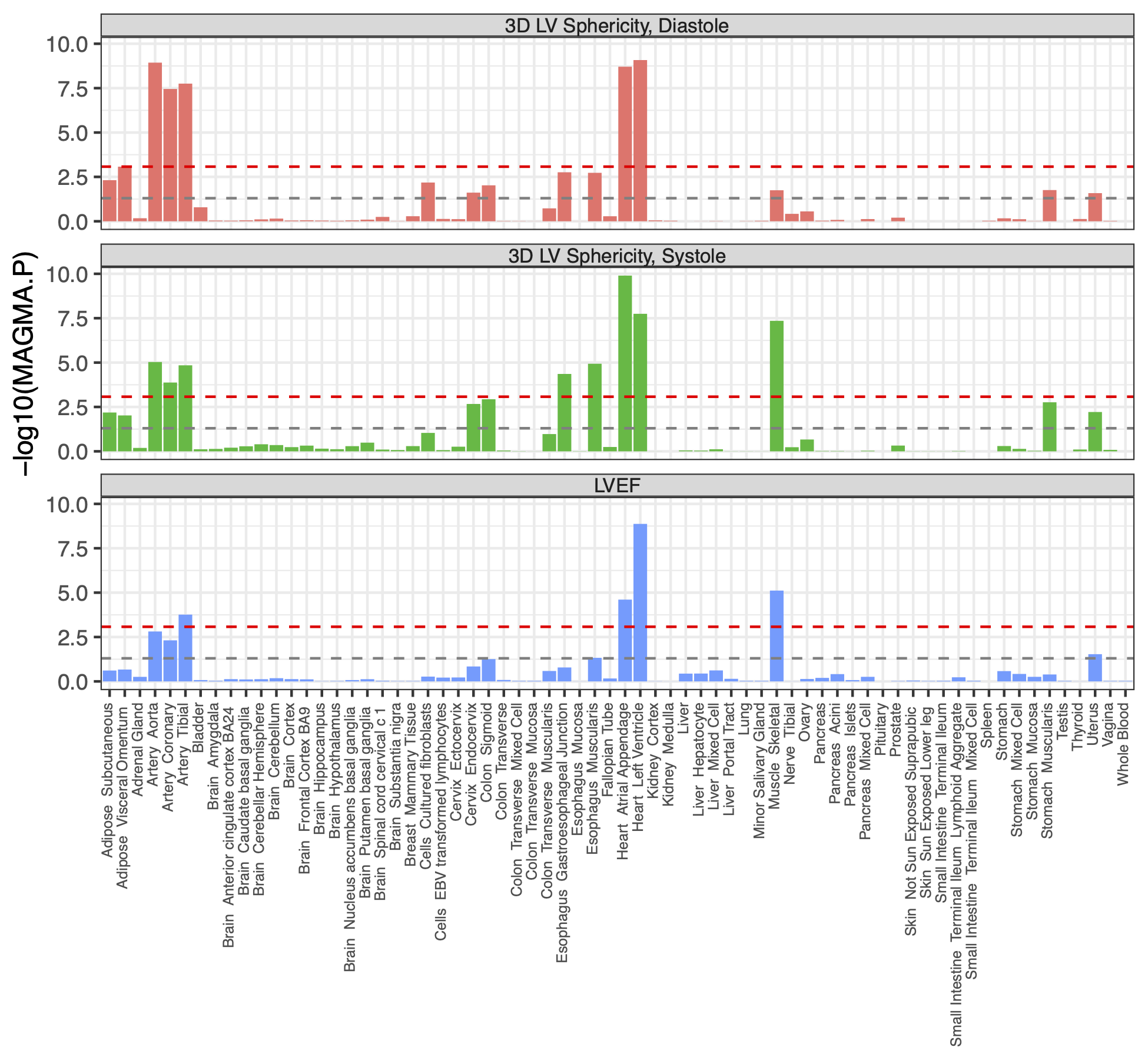

Statistical evidence for MAGMA gene-set overlap with GTEx v10 tissue types is depicted for LV sphericity and LVEF^32–34^.

#### Supplementary Figure 11: Single nucleus sequencing cell types from diseased hearts

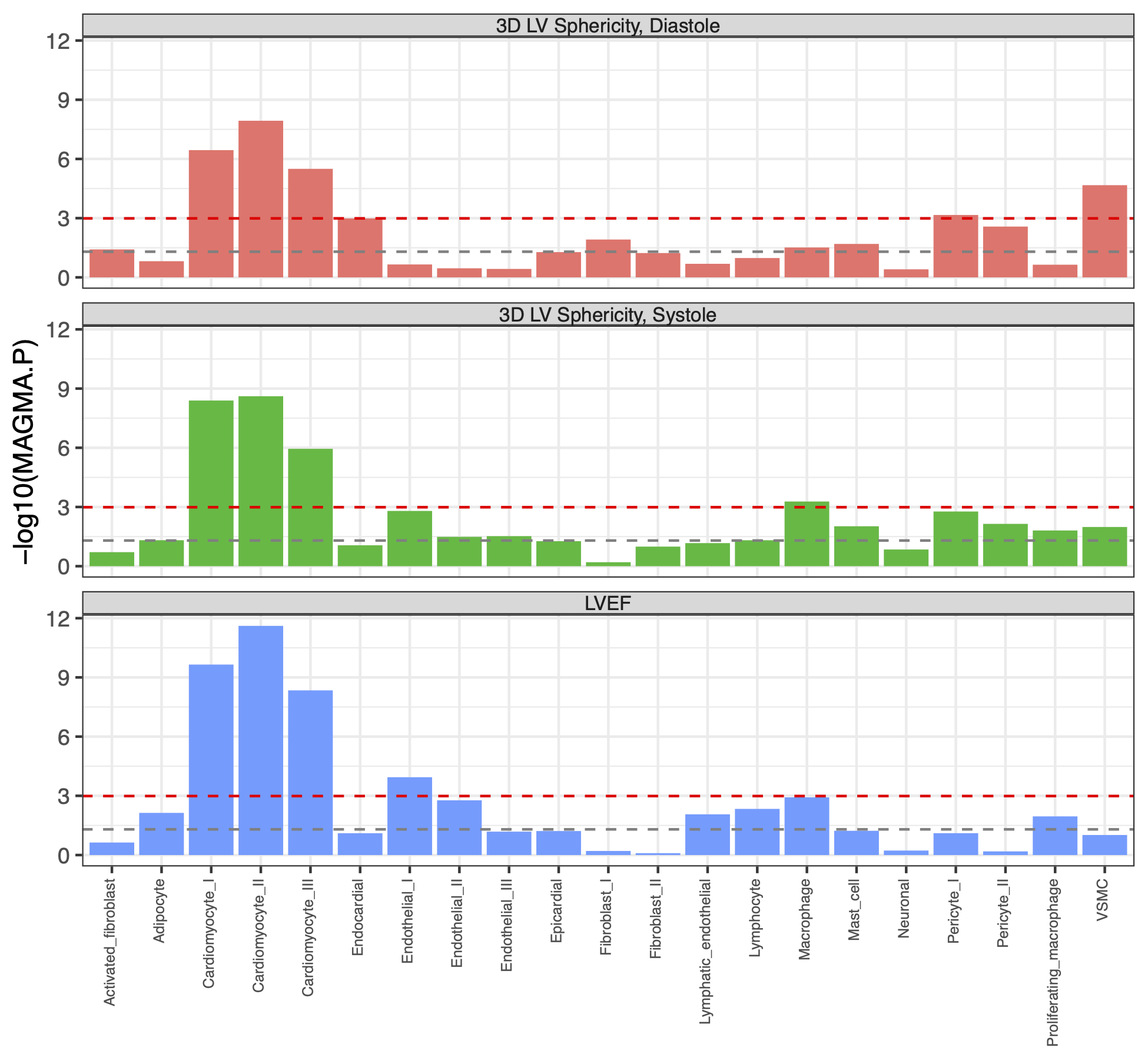

Statistical evidence for MAGMA gene-set overlap with single nucleus sequencing gene sets from Chaffin, *et al*, for LV sphericity and LVEF^35^.

#### Supplementary Figure 12: Single nucleus sequencing cell types from healthy hearts

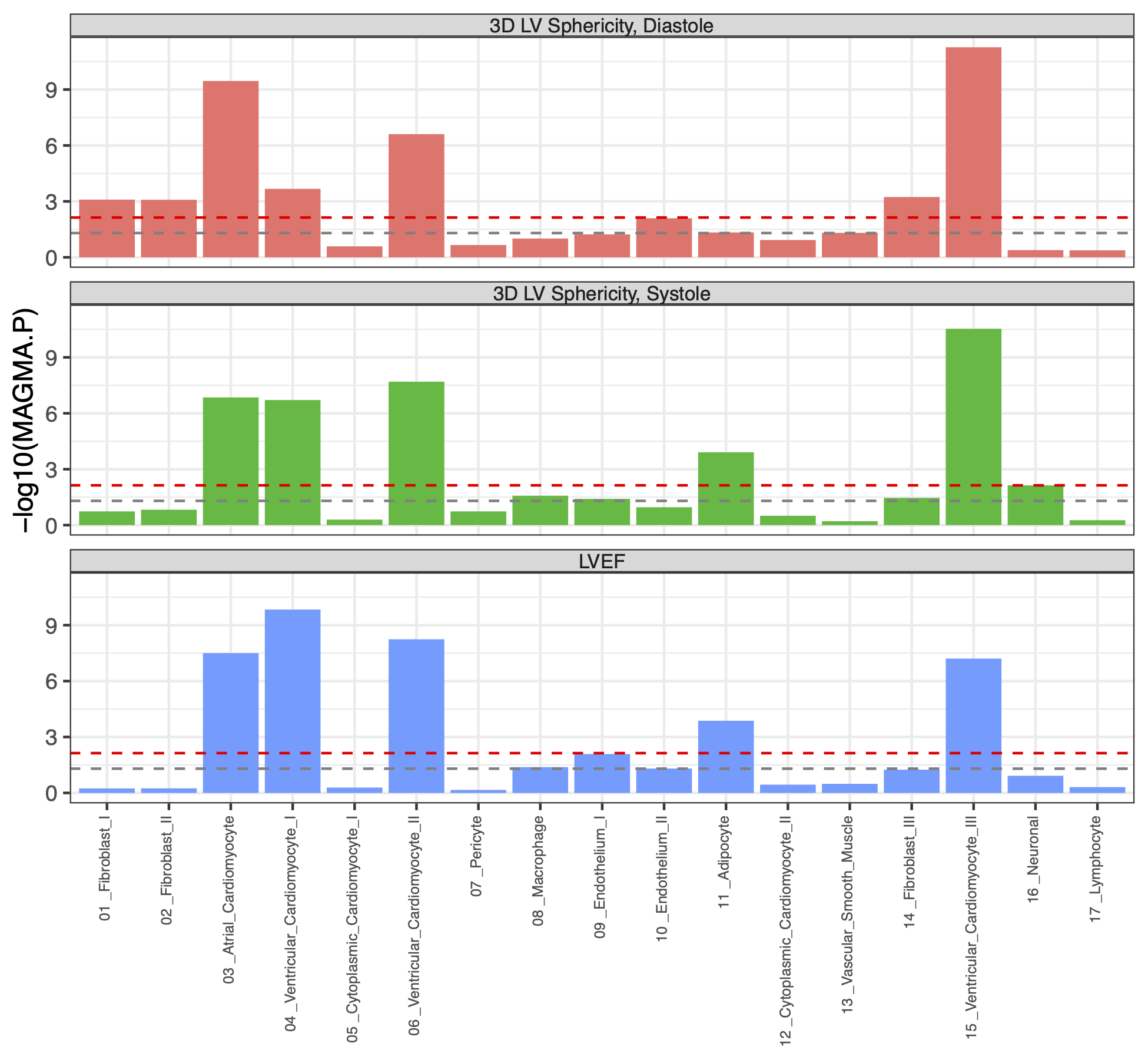

Statistical evidence for MAGMA gene-set overlap with single nucleus sequencing gene sets from Tucker, *et al*, for LV sphericity and LVEF^36^.

#### Supplementary Figure 13: Mendelian randomization volcano plots

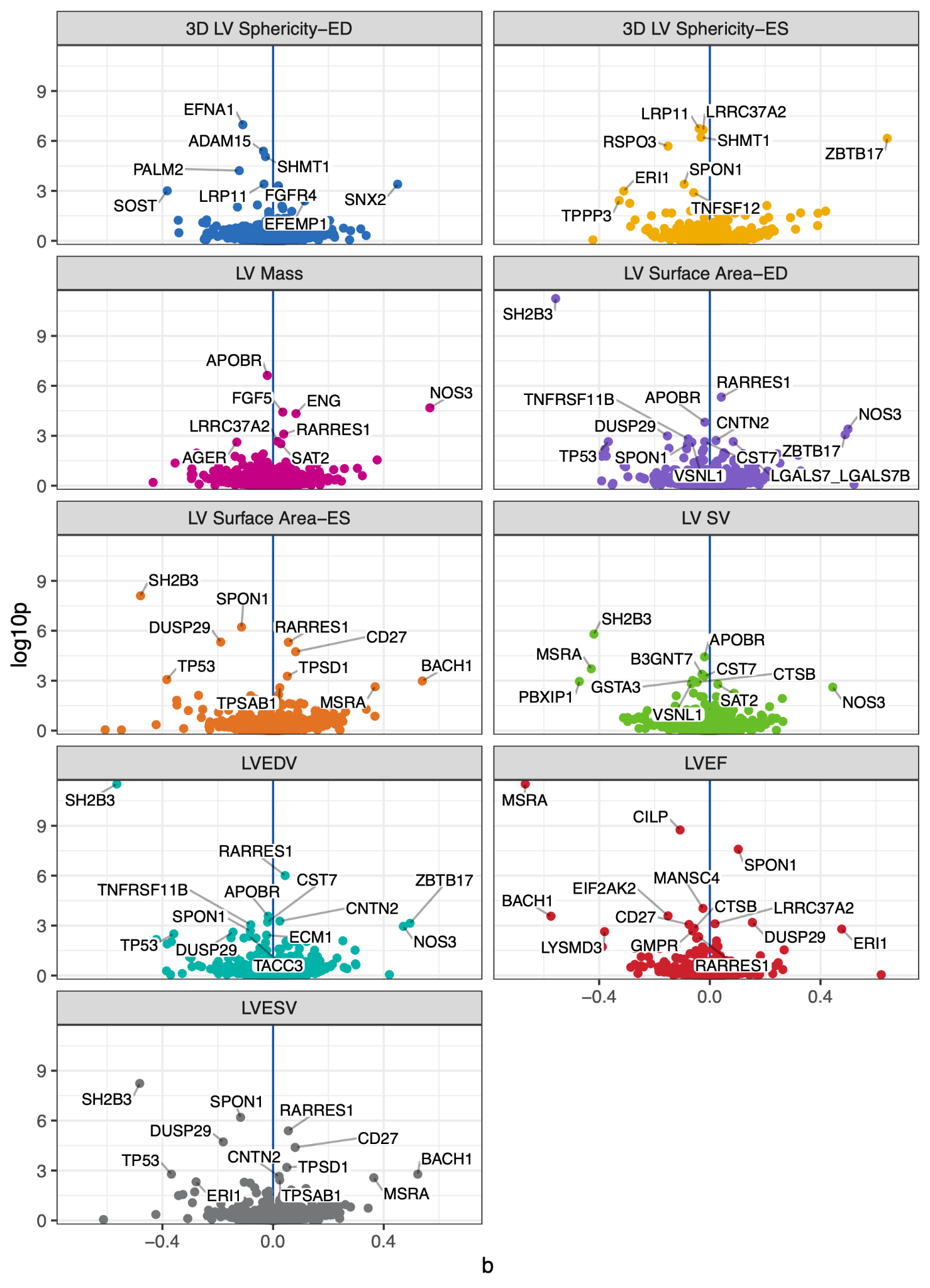

Volcano plots showing the causal effect estimate from Mendelian randomization of genetic dosage of Olink-measured proteins on LV measurements. The **x axis** represents the effect estimate, and the **y axis** represents the -log10 of the Benjamini-Hochberg adjusted P values. To allow visualization of the majority of the data, values with -log10(P) > 15 have been removed (which excludes ZBTB17 for LVEF, LVESV, and LV surface area-ES). To enhance readability, labels are not applied to all proteins with FDR < 0.05, but rather only to proteins with adjusted P < 0.005.

#### Supplementary Figure 14: Cumulative incidence of DCM with TTNtv in *All of Us*

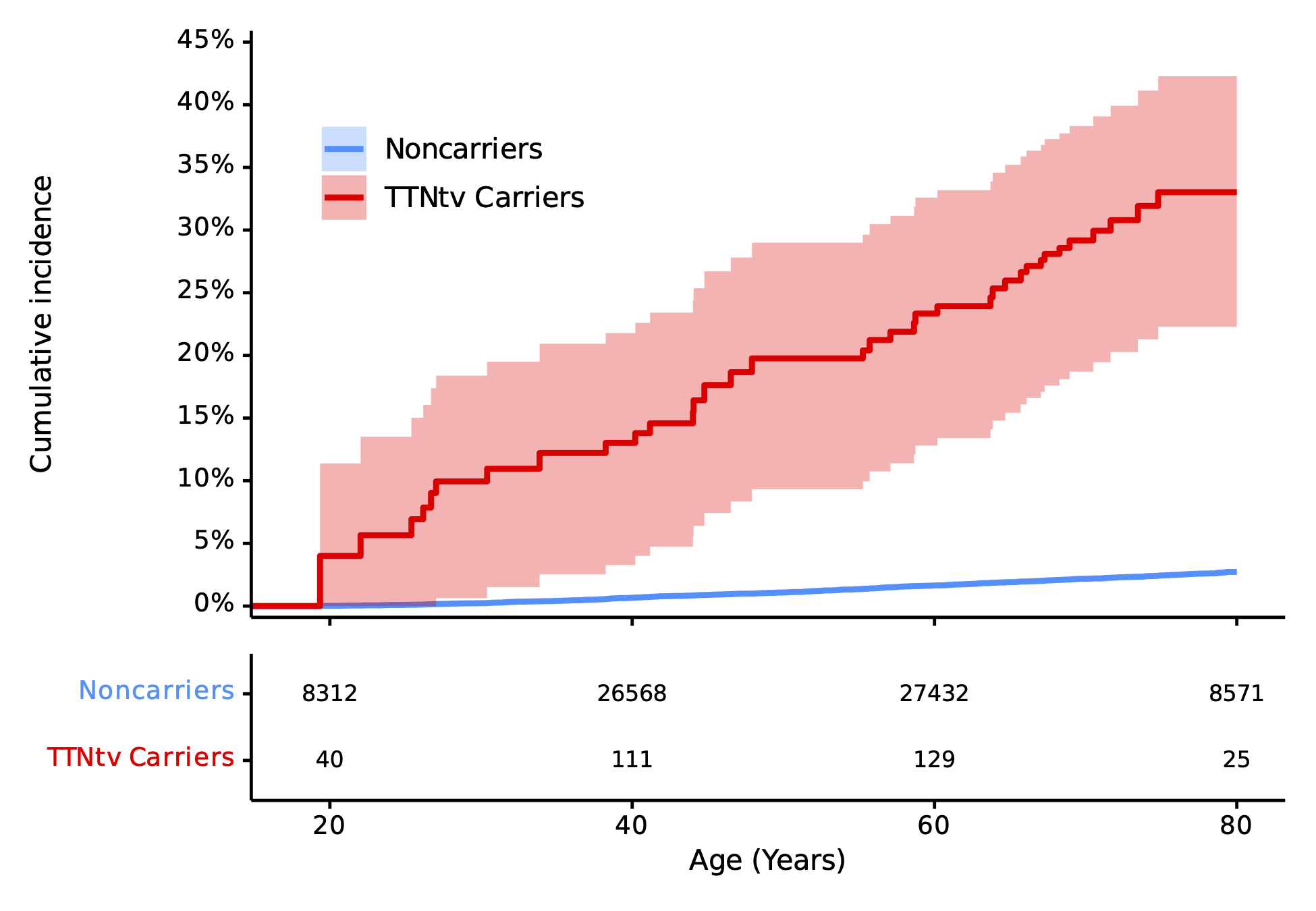

The Kaplan-Meier (KM) cumulative incidence for individuals in *All of Us* without DCM by age 18. Curves are left-truncated at each participant’s enrollment age, and therefore depict risk conditional on being alive, enrolled, disease-free, and uncensored at that attained age. The KM cumulative incidence by age 80 for TTNtv carriers was 33% (95% CI 22-42%). The mean duration of follow-up for participants was 3.6±1.7 years.
